# The Global Landscape of Genetic Variation in Parkinson’s disease: Multi-Ancestry Insights into Established Disease Genes and their Translational Relevance

**DOI:** 10.1101/2025.07.08.25330815

**Authors:** Lara M. Lange, Zih-Hua Fang, Mary B. Makarious, Nicole Kuznetsov, Kajsa Atterling Brolin, Shannon Ballard, Soraya Bardien, Maria Leila Doquenia, Peter Heutink, Henry Houlden, Hirotaka Iwaki, Simona Jasaityte, Lietsel Jones, Johanna Junker, Rauan Kaiyrzhanov, Mathew J. Koretsky, Kishore R. Kumar, the Latin American Research Consortium on the Genetics of Parkinson’s Disease (LARGE-PD), Hampton L. Leonard, Kristin S. Levine, Shen-Yang Lim, Niccoló E. Mencacci, Wael M. Y. Mohamed, Mike A. Nalls, Alastair J. Noyce, Rajeev Ojha, Njideka U. Okubadejo, Shoaib ur Rehman, Laurel Screven, Chingiz Shashkin, Sophia Sopromadze, Eleanor J. Stafford, Ai Huey Tan, Manuela Tan, Zaruhi Tavadyan, Joanne Trinh, Bayasgalan Tserensodnom, Enza Maria Valente, Dan Vitale, Nazira Zharkinbekova, Katja Lohmann, Sara Bandres-Ciga, Cornelis Blauwendraat, Andrew Singleton, Huw R. Morris, Christine Klein, the Global Parkinson’s Genetics Program (GP2)

## Abstract

**Background:** The genetic architecture of Parkinson’s disease (PD) varies considerably across ancestries, yet most genetic studies have focused on individuals of European descent, limiting insights into the genetic landscape of PD at a global scale.

**Methods:** We conducted a large-scale, multi-ancestry investigation of causal and risk variants in PD-related genes. Using genetic datasets from the Global Parkinson’s Genetics Program (GP2), we analyzed sequencing and genotyping data from 105,588 individuals, including 63,837 affected and 41,751 unaffected, from eleven different ancestries. Approximately 29% of individuals included were from underrepresented populations.

**Findings:** Our findings revealed shared and ancestry-specific patterns in the prevalence and variant spectrum. Overall, ∼2% of PD individuals carried a causative variant, with substantial variations across ancestries ranging from ∼0·5% in African to ∼7% in Middle Eastern and >10% Ashkenazi Jewish ancestries. Including disease-associated *GBA1* and *LRRK2* risk variants raised the yield to 13·6%, largely driven by *GBA1*, except in East Asians, where *LRRK2* risk variants dominated. *GBA1* variants were most frequent globally, albeit with substantial differences in frequencies and variant spectra. While *GBA1* variants were identified across all ancestries, frequencies ranged from ∼5% in Middle Eastern and South Asian to ∼52% in African ancestry. Similarly, *LRRK2* variants showed ancestry-specific enrichment, with p.G2019S most frequently seen in Middle Eastern and Ashkenazi Jewish populations, and risk variants predominating in East Asians. Notably, clinical trials targeting specific genetic variants are currently primarily based in Europe and North America. Another globally PD-relevant gene was *PRKN*, with variant carriers identified across almost all ancestries.

**Interpretation:** This large-scale, multi-ancestry assessment offers crucial insights into the population-specific genetic architecture of PD. It underscores the critical need for increased ancestral diversity in PD research to improve diagnostic accuracy, enhance our understanding of disease mechanisms across populations, and ensure equitable development and application of emerging precision therapies.

**Funding:** Aligning Science Across Parkinson’s (ASAP) Global Parkinson’s Genetics Program (GP2)

## Evidence before this study

Genetic discoveries have dramatically advanced our understanding of Parkinson’s disease (PD), including the identification of rare monogenic causal and common risk variants. However, according to our PubMed-based (https://pubmed.ncbi.nlm.nih.gov/) literature review, the majority of genetic data has been generated in PD cohorts of European ancestry. The sample sets of two recent comprehensive, large-scale genetic screening studies, the PD GENEration North America and the Rostock International Parkinson’s disease (ROPAD) study, comprised 85% and 96% of White participants, respectively. Moreover, a global survey of monogenic PD, published in 2023, reported that 91% of genetically confirmed individuals with PD were of European ancestry, underscoring the major gap in ancestral representation in PD genetics research. Although ancestry-specific variants and founder effects have been reported for select PD-associated genes such as *GBA1*, *LRRK2*, and *PINK1*, comprehensive, globally- and ancestry-representative studies are missing, thereby constraining the applicability of genetic diagnostics and the development and inclusivity of equitable, ancestry-informed therapeutic strategies.

### Added value of this study

This study, conducted within the framework of the Global Parkinson’s Genetics Program (GP2), represents the largest multi-ancestry exploratory genetic investigation of PD to date, including over 105,000 individuals from around the world, with more than 30,000 participants of non-European and non-Ashkenazi Jewish ancestry (representing ∼29% of the study cohort). Importantly, this includes the largest systematically curated genetic resource currently available for multiple historically underrepresented populations, for whom sample collection and data curation are logistically challenging and genetic reference data remain sparse. Even where absolute sample sizes are much smaller than in European cohorts, these data are informative and of high translational value, providing essential context for variant interpretation and ancestry-specific disease biology.

By exploring pathogenic variants in established PD-linked genes and PD risk-associated variants across 11 diverse genetically defined ancestry groups, this study provides a comprehensive global view of the genetic architecture of PD. This work highlights both shared and ancestry-specific contributions, with clear clinical and translational implications.

Variability in *GBA1* is a globally relevant genetic contributor to PD risk, with variant carriers identified across all investigated populations, reinforcing its relevance as a therapeutic target across ancestries. Similarly, *LRRK2* variant carriers, currently already targeted in clinical trials, and individuals with *PRKN* alterations, increasingly being explored as a therapeutic target in preclinical studies, were identified across multiple ancestral groups, supporting the broader applicability of genetically targeted approaches.

At the same time, substantially different genetic yields across ancestries underscore the limited transferability of current PD gene and variant panels, which are largely derived from European-ancestry datasets. This highlights the need for ancestry-aware interpretation frameworks and illustrates how variants identified in underrepresented populations often remain of uncertain significance due to limited comparative data. By addressing these gaps, this study establishes a meaningful reference framework and a foundation for future ancestry-specific analyses, functional studies, and clinically actionable genetic research.

### Implications of all the available evidence

As precision medicine approaches are increasingly integrated into PD research and clinical trials, it is critical to ensure that emerging therapies are applicable and accessible to individuals of all ancestral backgrounds. Current genetically stratified trials predominantly enroll participants of European or Ashkenazi Jewish ancestry and trial sites are predominantly located across Europe and North America, raising concerns about equity, generalisability, and global clinical impact. This study underscores the importance of globally inclusive genetic efforts such as GP2 to close these existing gaps, improve diagnostic equity, inform ancestry-aware trial design, and enable more representative and effective implementation of precision neurology worldwide.

## Introduction

Genetic discoveries have transformed our understanding of Parkinson’s disease (PD). Genetic factors substantially contribute to PD risk and progression,^1^ comprising a spectrum ranging from rare variants with large effect sizes to common variants with small effect sizes. More than 130 independent risk loci have been identified by genome-wide association studies,^2^ and over 15 genes have been linked to monogenic PD and parkinsonism.^1,3^

A critical gap in PD genetics research is the lack of ancestral diversity.^4^ Although genetic forms of PD are found globally, approximately 75% of all genetic PD studies to date focused on European-ancestry individuals.^5,6^ Research in diverse populations is critical to uncovering unique genetic contributions to disease and improve diagnosis, risk assessment, and genetic counseling. This is particularly relevant for clinical trials driven by genetic discoveries, e.g., targeting glucocerebrosidase or LRRK2 kinase (encoded by the PD-linked genes *GBA1* and *LRRK2*, respectively). Lack of diversity may limit the global applicability of emerging opportunities, as it is unclear whether current genetic targets are relevant across ancestries or whether population-specific variants exist. Expanding genetic studies to all ancestries is essential to improve diagnostic accuracy and therapeutic development.

The Global Parkinson’s Genetics Program (GP2; https://gp2.org/)^7^ is a large-scale initiative actively addressing this gap by investigating the genetic underpinnings of PD and parkinsonism across global populations.^5,8^ The aim of this study was to investigate pathogenic and high-risk variants linked to PD and parkinsonism at a global scale using data from GP2.

## Materials and Methods

### Study design and participants

Our study workflow is displayed in Figure 1. We analyzed short-read genome sequencing, clinical exome sequencing and Illumina NeuroBooster Array genotyping data from GP2 Release 11 (https://doi.org/10.5281/zenodo.17753486). A detailed sample inventory is provided in Supplementary Table 1. The total sample comprised 105,588 individuals, including 58,559 individuals with PD, 5,278 with other neurodegenerative or movement disorder phenotypes (atypical or unspecified parkinsonism, essential tremor, Alzheimer’s disease and other dementias), 512 unaffected family members of those individuals, and 41,239 controls. Data processing, quality control, and genetic ancestry prediction were performed with the genotools pipeline, as described elsewhere.^9^ Samples were from eleven different ancestries: African-Admixed (AAC), African (AFR), Ashkenazi Jewish (AJ), Latinos and Indigenous People of the Americas (AMR), Complex Admixture (CAH), Central Asian (CAS), East Asian (EAS), European (EUR), Finnish (FIN), Middle Eastern (MDE), and South Asian (SAS). Because the CAH group comprises highly admixed individuals for whom ancestry-specific interpretation is limited, this group is not discussed in the Results, but all corresponding carrier numbers are provided in the (Supplementary) Tables.

**Figure 1.**
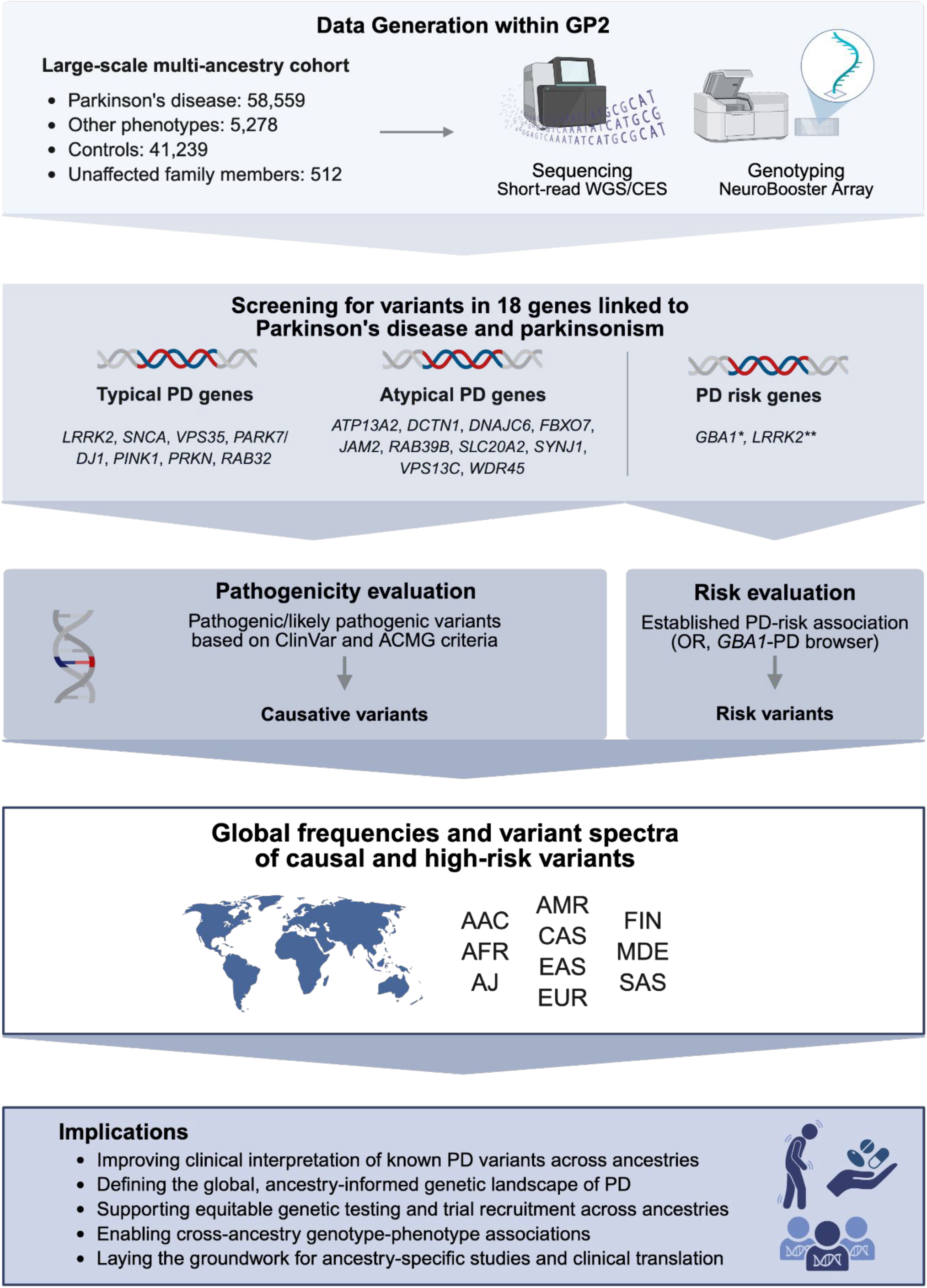
Study Workflow and Implications. Individuals were screened for causative (pathogenic/likely pathogenic according to ClinVar and/or ACMG criteria) and selected risk-associated variants in 18 distinct PD-linked genes using short-read sequencing and genome-wide genotyping data. *For the purpose of this study and in the context of PD, all variants in GBA1 (including variants causal for Gaucher’s disease) are considered PD risk variants (regardless of their Gaucher’s severity). **Select *LRRK2* variants with established PD-risk associations included rs33949390 (chr12:40320043:G:C, R1628P) and rs34778348 (chr12:40363526:G:A, G2385R). Figure created with BioRender. AAC = African Admixed, ACMG = American College of Medical Genetics and Genomics, AFR = African, AJ = Ashkenazi Jewish, AMR = Latinos and Indigenous people of the Americas, CAH = Complex Admixture, CES = clinical exome sequencing, CAS = Central Asian, EAS = East Asian, EUR = European, FIN = Finnish, GP2 = Global Parkinson’s Genetics Program, MDE = Middle Eastern, OR = Odds ratio, PD = Parkinson’s disease, SAS = South Asian, WGS = whole-genome sequencing.

### Genes and variants of interest

We focused on variants in genes linked to PD and parkinsonism following the recommendations of the *MDS Task Force on the Nomenclature of Genetic Movement Disorders*^3^, including variants in i) *LRRK2, SNCA,* and *VPS35* linked to autosomal-dominant PD, ii) *DJ-1/PARK7, PINK1,* and *PRKN* linked to autosomal-recessive PD, and iii) *ATP13A2*, *DCTN1, DNAJC6, FBXO7, JAM2, RAB39B, SLC20A2, SYNJ1, VPS13C,* and *WDR45* linked to atypical or complex parkinsonism. We included only PARK-designated genes; genes linked to combined phenotypes including PD or parkinsonism (e.g., dystonia-parkinsonism, Frontotemporal Dementia-parkinsonism, etc.) were beyond the scope of this study. We added *RAB32* p.S71R, identified as causal for PD after the Task Force recommendations were published. We included pathogenic/likely pathogenic (according to ClinVar; https://www.ncbi.nlm.nih.gov/clinvar/) variants in these genes and also investigated PD risk-associated variants in *GBA1* and *LRRK2*, given their established translational significance. For details on pathogenicity evaluation, see Supplementary Material. Copy number variation (CNV) analyses for *SNCA* and *PRKN* were performed for all samples with available genotyping data as described before.^10^ Carriers of two pathogenic variants in recessive genes and an age at onset (AAO) ≤60 years were considered likely compound heterozygous without further validation, based on findings from previous studies.^11^

## Results

### Summary of global genetic findings in GP2

Sample characteristics are summarized in Table 1. Across all ancestries, 2·1% (1217/58559) of individuals with PD carried a causative and 11·8% (6893/58559) a risk variant, whereas yields in individuals with other neurodegenerative or movement disorder phenotypes were substantially lower with 0.3% (16/5278) harboring causative and 6.7% (355/5278) harboring risk variants (Table 2). In comparison, causative variants were observed in 0.3% (106/41239) and risk variants in 8·7% (3585/41239) of controls (Table 2).

**Table 1.** Study sample characteristics. Summary of sample numbers and cohort-level data by study type, including sex, age, age at onset (if applicable), and family history of Parkinson’s disease.

| Group | Samples (n) | Male (%) | Median Age* (range) | Median AAO** (range) | Positive FH of PD*** (%) |
| --- | --- | --- | --- | --- | --- |
| **Affected individuals** | 63837 | 39193 (61.4) | 67 (13-100),<br>[unk for n = 11969] | 60 (10-98),<br>[unk for n = 16623] | 11821 (18.5),<br>[unk for n = 23860] |
| PD | 58559 | 36178 (61.8) | 67 (13-100),<br>[unk for n = 8437] | 60 (10-98),<br>[unk for n = 13791] | 11543 (19.7),<br>[unk for n = 19893] |
| Other phenotypes | 5278 | 3015 (57.1) | 70 (22-99),<br>[unk for n = 3532] | 65 (10-88),<br>[unk for n = 2832] | 278 (5.3),<br>[unk for n = 3967] |
| **Unaffected individuals** | 41751 | 20717 (49.6) | 63 (10-99),<br>[unk for n = 7614] | NA | 1678 (4.0),<br>[unk for n = 20193] |
| Controls | 41239 | 20491 (49.7) | 63 (10-99),<br>[unk for n = 7576] | NA | 1166 (4.9),<br>[unk for n = 28059] |
| Unaffected family members | 512 | 226 (44.1) | 53 (12-92),<br>[unk for n = 38] | NA | NA |
| **TOTAL** | **105588** | **59910 (56.7)** | **66 (10-100),**<br>**[unk for n = 19583]** | **NA** | **13499 (12.8),**<br>**[unk for n = 51919]** |

\*Age refers to the age at recruitment/age at sample collection; individuals with an age <10 years or >100 years were considered outliers and excluded from these statistics.
\*\* Individuals with an AAO <10 years or >100 years were considered outliers and excluded from these statistics. \*\*\* Including related individuals.
AAO = Age at onset, FH = Family history, NA = not available/not applicable, PD = Parkinson's disease, unk = unknown, PD = Samples from studies recruiting PD cases based on different selection criteria (i.e., sporadic PD, early-onset or familial PD, case-control studies, etc.). This group only includes individuals with a PD diagnosis. Other phenotypes = Samples from individuals with diagnoses other than PD, including atypical parkinsonism (e.g., Dementia with Lewy Bodies [DLB], Corticobasal Degeneration/Syndrome [CBD/CBS], Multisystem Atrophy [MSA], and Progressive Supranuclear Palsy [PSP]), unspecified parkinsonism, Essential Tremor, and Alzheimer's disease and other dementias. Controls = Neurologically healthy controls. Unaffected family members = Individuals that are unaffected family members of individuals with PD.

**Table 2.** Summary of genetic findings and yields across ancestries. Proportion of individuals with genetic findings identified in this study, stratified by phenotype, ancestry and variant type.

| Phenotype | Variant category | AAC | AFR | AJ | AMR | CAH | CAS | EAS | EUR | FIN | MDE | SAS | Total |
| --- | --- | --- | --- | --- | --- | --- | --- | --- | --- | --- | --- | --- | --- |
| PD | Causal variants (yield, %) | 3/550 (0.5) | 10/2844 (0.4) | 251/2343 (10.7) | 85/2809 (3.0) | 35/902 (3.9) | 9/1284 (0.7) | 69/4773 (1.44) | 651/40773 (1.6) | 1/127 (0.8) | 92/1347 (6.8) | 11/807 (1.4) | 1217/58559 (2.1) |
|  | Risk variants (yield, %) | 198/550 (36.0) | 1507/2844 (53.0) | 394/2343 (16.8) | 162/2809 (5.8) | 148/902 (16.4) | 98/1284 (7.6) | 796/4773 (16.7) | 3452/40773 (8.5) | 26/127 (20.5) | 73/1347 (5.4) | 39/807 (4.8) | 6893/58559 (11.8) |
|  | **Total (yield, %)** | **199/550 (36.2)** | **1510/2844 (53.1)** | **620/2343 (26.5)** | **245/2809 (8.7)** | **179/902 (19.8)** | **105/1284 (8.2)** | **840/4773 (17.6)** | **4054/40773 (9.9)** | **26/127 (20.5)** | **158/1347 (11.7)** | **49/807 (6.1)** | **7985/58559 (13.6)** |
| Other | Causal variants (yield, %) | 0/11 (0) | 0/43 (0) | 2/98 (2.1) | 2/26 (7.7) | 0/43 (0) | 0/131 (0) | 1/662 (0.2) | 10/4153 (0.2) | 0/9 (0) | 1/28 (3.6) | 0/74 (0) | 16/5278 (0.3) |
|  | Risk variants (yield, %) | 2/11 (18.2) | 14/43 (32.6) | 15/98 (15.8) | 0/26 (0) | 3/43 (7.0) | 2/131 (1.5) | 60/662 (9.1) | 253/4153 (6.1) | 3/9 (33.3) | 2/28 (7.1) | 1/74 (1.4) | 355/5278 (6.7) |
|  | **Total (yield, %)** | **2/11 (18.2)** | **14/43 (32.6)** | **16/98 (16.3)** | **2/26 (7.7)** | **3/43 (7.0)** | **2/131 (1.5)** | **60/662 (9.1)** | **263/4153 (6.3)** | **3/9 (33.3)** | **3/28 (10.7)** | **1/74 (1.4)** | **369/5278 (7.0)** |
| Controls | Causal variants (yield, %) | 3/881 (0.3) | 4/4495 (0.1) | 20/913 (2.2) | 0/1654 (0) | 2/444 (0.5) | 1/1480 (0.1) | 2/2809 (0.1) | 69/26761 (0.3) | 0/22 (0) | 5/1340 (0.4) | 0/440 (0) | 106/41239 (0.3) |
|  | Risk variants (yield, %) | 211/881 (24.0) | 1692/4495 (37.6) | 76/913 (8.3) | 30/1654 (1.8) | 46/444 (10.4) | 43/1480 (2.9) | 185/2809 (6.6) | 1274/26761 (4.8) | 4/22 (18.2) | 20/1340 (1.5) | 4/440 (0.9) | 3585/41239 (8.7) |
|  | **Total (yield, %)** | **213/881 (24.2)** | **1693/4495 (37.7)** | **95/913 (10.4)** | **30/1654 (1.8)** | **48/444 (10.8)** | **44/1480 (3.0)** | **187/2809 (6.7)** | **1337/26761 (5.0)** | **4/22 (18.2)** | **25/1340 (1.9)** | **4/440 (0.9)** | **3680/41239 (8.9)** |
| Healthy family members | Causal variants (yield, %) | 1/3 (33.3) | 0/1 (0) | 2/6 (33.3) | 0/2 (0) | 3/6 (50) | 0/66 (0) | 0/80 (0) | 4/288 (1.4) | NA | 0/6 (0) | 0/54 (0) | 10/512 (2.0) |
|  | Risk variants (yield, %) | 0/3 (0) | 0/1 (0) | 0/6 (0) | 1/2 (50) | 0/6 (0) | 1/66 (1.5) | 12/80 (15.0) | 25/288 (8.7) | NA | 0/6 (0) | 5/54 (9.3) | 44/512 (8.6) |
|  | **Total**<br>**(yield, %)** | **1/3**<br>**(33.3)** | **0/1**<br>**(0)** | **2/6**<br>**(33.3)** | **1/2**<br>**(50)** | **3/6**<br>**(50)** | **1/66**<br>**(1.5)** | **12/80**<br>**(15.0)** | **29/288**<br>**(10.1)** | **NA** | **0/6**<br>**(0)** | **5/54**<br>**(9.3)** | **54/512**<br>**(10.5)** |

This table summarizes counts and proportions of individuals carrying risk variants only, causal variants only, or both. The combined category represents unique individuals with any qualifying variant. Because some individuals carry both risk and causal variants, combined counts and percentages are lower than the sum of category-specific values.
AAC = African Admixed, AFR = African, AJ = Ashkenazi Jewish, AMR = Latino and Indigenous people of the Americas, CAH = Complex Admixture, CAS = Central Asian, EAS = East Asian, EUR = European, FIN = Finnish, MDE = Middle Eastern, SAS = South Asian

Across all affected variant carriers, we identified 117 distinct pathogenic single nucleotide variants (SNVs) in 15 genes (including single heterozygous variants in recessive genes), alongside two *LRRK2* risk variants, 99 *GBA1* variants, and structural variations like *SNCA* multiplications and *PRKN* exon deletions and duplications.

The global genetic spectrum of PD across ancestries is summarised in Figure 2, Supplementary Figure 1 and Supplementary Table 2, illustrating both shared genetic contributors and marked ancestry-specific distributions. *GBA1* variant carriers were most frequent overall and detected across all ancestries, albeit with substantially different variant spectra (Figure 3). Similarly, *LRRK2* variants were identified across multiple ancestries with ancestry-specific variant profiles (Figure 4). Rarer *PRKN* SNVs and structural variations were also observed across most ancestries, whereas causative variants in other genes were only identified in certain populations. Particularly, variants in genes linked to atypical parkinsonism were only identified in a small subset of individuals (Supplementary Table 3). Individuals with *GBA1*-associated PD showed a significantly earlier median AAO of 3 to 8 years across multiple ancestries compared to those with idiopathic PD (IPD) (Supplementary Figure 2A and Supplementary Table 4). The AAO between *LRRK2*-PD and IPD showed similar trends in selected ancestries, with *LRRK2*-PD showing a 1- to 3-year earlier onset (Supplementary Figure 2A and Supplementary Table 4). Further, the AAO distribution among *GBA1* and *LRRK2* carriers showed heterogeneity across ancestries (Supplementary Figures 2B and C). Detailed results for each ancestry are reported below.

**Figure 2.**
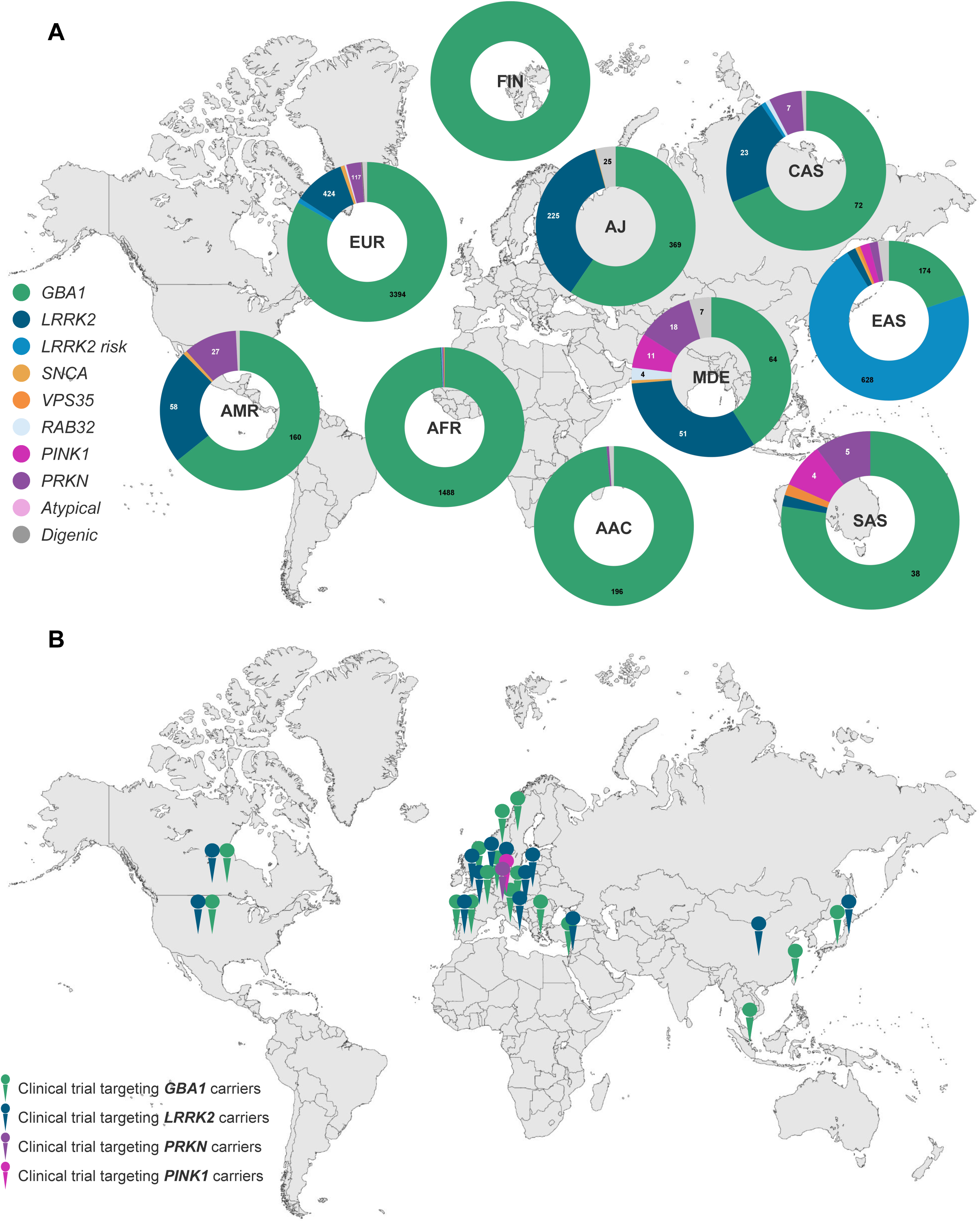
Global genetic spectrum of PD and locations of clinical trials targeting gene variant carriers. (A) Donut charts displaying the percentage of identified carriers per gene amongst all carriers per ancestry. Dual refers to carriers of variants in two different genes (e.g., *GBA1* and a causal gene). (B) Global map of clinical trial locations recruiting gene variant carriers (*GBA1, LRRK2, PINK1,* and *PRKN*) and targeting proteins encoded by PD-linked genes or pathways in which these genes are involved, obtained from the clinical trial registry https://clinicaltrials.gov/. Only one pin per gene and country is shown, regardless of the number of study sites within that country. AAC = African Admixed, AFR = African, AJ = Ashkenazi Jewish, AMR = Latinos and Indigenous people of the Americas, CAS = Central Asian, EAS = East Asian, EUR = European, FIN = Finnish, MDE = Middle Eastern, SAS = South Asian.

**Figure 3.**
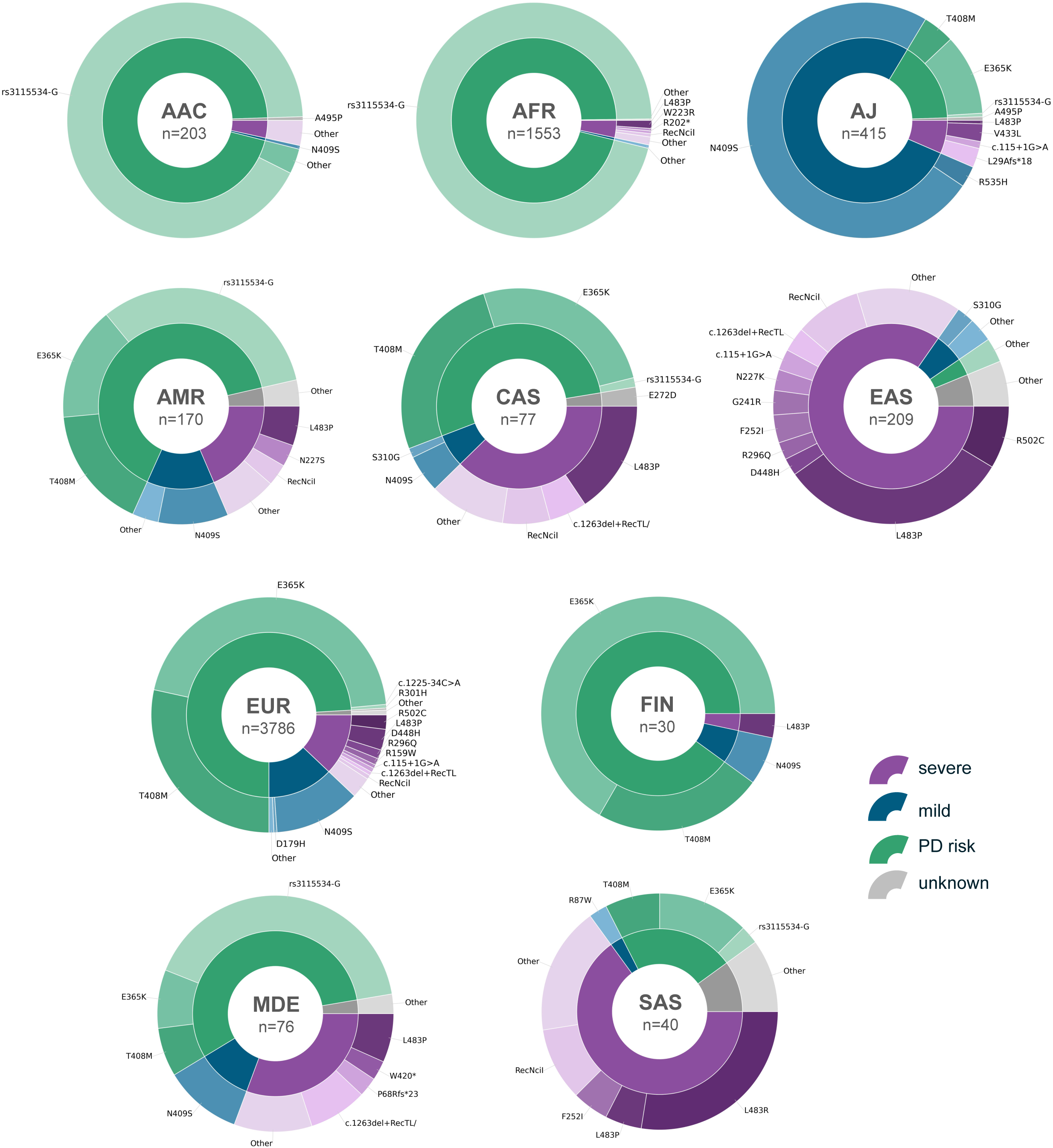
Mutational and severity spectrum of *GBA1* variants across ancestries. Donut charts illustrate the distribution of *GBA1* variants in individuals with Parkinson’s disease and other neurodegenerative or movement disorder phenotypes stratified by variant severity (inner ring: Severe, Mild, Risk, Unknown) and specific variants (outer ring) across ancestries. Variant severity was assigned using the GBA1-PD browser (https://pdgenetics.shinyapps.io/GBA1Browser/). The size of each segment reflects the number of variant carriers within each ancestry. Inner-ring colors denote severity categories: Severe (purple), Mild (blue), Risk (green), and Unknown (gray). Outer-ring segments represent individual variants, with color shades corresponding to their severity classification. To improve readability, variants with a single carrier were grouped as “Other” for CAS, MDE, and SAS; for AAC, AFR, AMR, EAS, and EUR, variants with ≤5 carriers were grouped. Numbers within the circles indicate the total number of *GBA1* variants identified per ancestry; individuals carrying two different *GBA1* variants contributed to both variant counts. AAC = African Admixed, AFR = African, AJ = Ashkenazi Jewish, AMR = Latinos and Indigenous people of the Americas, CAH = Complex Admixture, CAS = Central Asian, EAS = East Asian, EUR = European, FIN = Finnish, MDE = Middle Eastern, SAS = South Asian.

**Figure 4.**
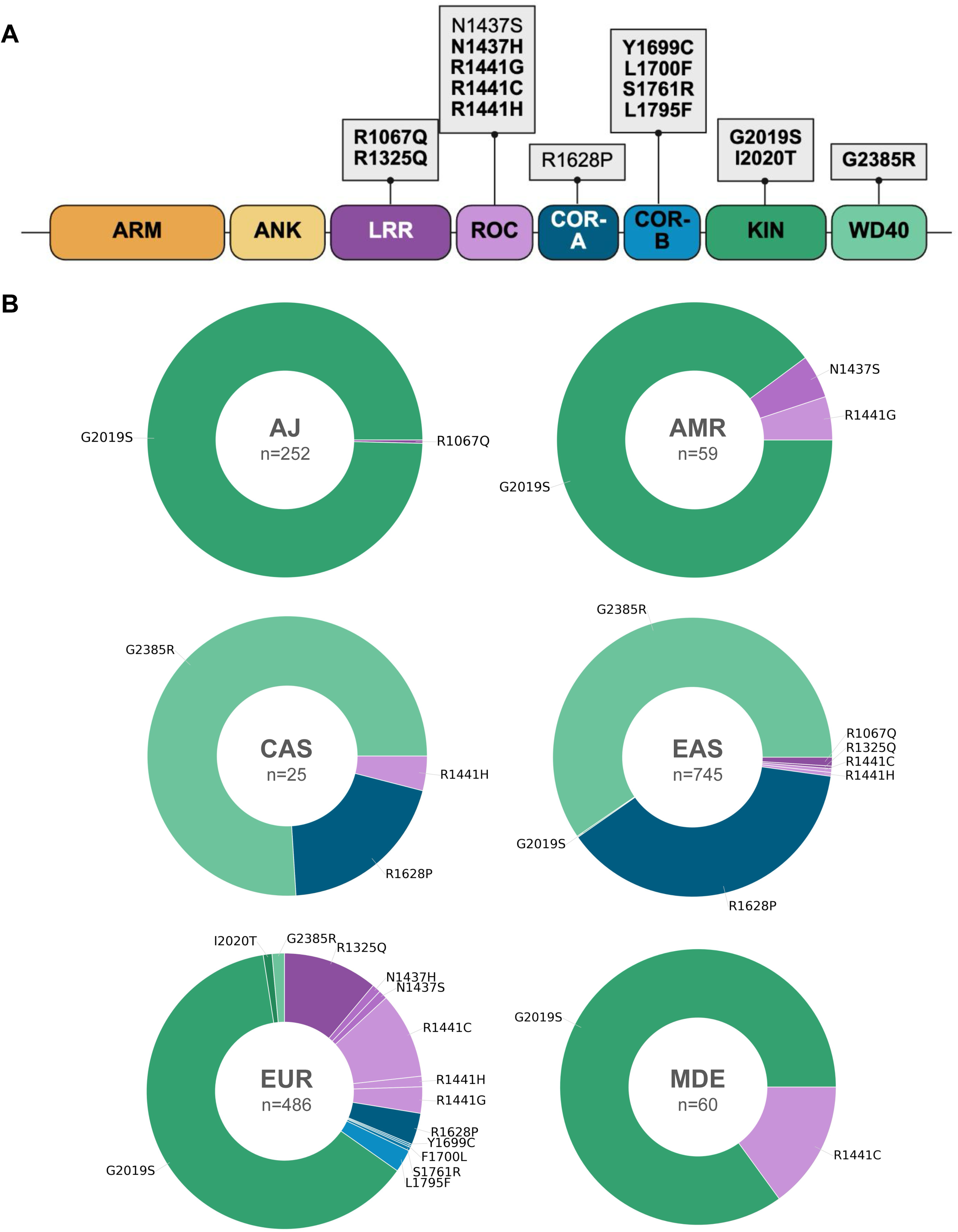
Distribution of *LRRK2* variants across protein domains and ancestries. **(A)** Schematic representation of the LRRK2 protein with functional domains. Boxes indicate locations of the identified *LRRK2* variants within their respective domains. Variants highlighted in bold have been previously shown to increase LRRK2 kinase activity. **(B)** Donut charts illustrate the ancestry-specific spectrum of *LRRK2* variants in individuals with Parkinson’s disease and other neurodegenerative and movement disorder phenotypes. Each segment represents a specific variant; the segment size is proportional to the number of individuals carrying that variant. If two variants were present in one individual, both variants were counted. Segment colors correspond to the LRRK2 protein domain in which the variant is located, as indicated in Panel A. Only ancestries with more than five variant carriers are shown. AJ = Ashkenazi Jewish, AMR = Latinos and Indigenous people of the Americas, CAS = Central Asian, EAS = East Asian, EUR = European, MDE = Middle East

### African-Admixed ancestry

Among individuals with PD, pathogenic or risk variants were identified in 36·2% (199/550). Almost all carried *GBA1* variants (198/199, 99·5%), predominantly driven by the common intronic risk variant, rs3115534-G (Figure 3), with both heterozygous and homozygous carriers. One individual (0·2%) with early-onset PD (AAO 27 years) carried a homozygous *PRKN* deletion, and another one (0·2%) harboured both a pathogenic *LRRK2* and a *GBA1* risk variant.

Among individuals with other neurological phenotypes, *GBA1* variants were identified in two individuals (2/11, 18·2%). In controls, *GBA1* variants were observed in 23·8% (210/881), again largely attributable to the rs3115534-G risk variant, predominantly in heterozygous state; pathogenic or risk *LRRK2* variants were identified in 0·5% (4/881).

### African ancestry

Pathogenic or risk variants were detected in 53·1% (1510/2844) of individuals with PD. *GBA1* variants accounted for the majority of genetic findings, again predominantly driven by rs3115534-G (Figure 3), with both heterozygous and homozygous carriers. Less frequent findings included variants in *LRRK2* (5/2844, 0·2%) and *SNCA* (1/2844, <0·1%), as well as variants in the recessive genes *PRKN* (4/2844, 0·1%) and *PINK1* (1/2844, <0·1%), observed in individuals with earlier AAO (ranging from 27-54 years); most of these individuals (7/12) carried a co-occurring *GBA1* variant.

Among individuals with other neurological phenotypes, only variants in *GBA1* were observed (14/43, 32·6%). In controls, genetic findings were identified in 37·7% (1693/4495), predominantly in *GBA1*, largely attributable to rs3115534-G, with pathogenic or risk *LRRK2* variants observed in a few individuals (4/4495, <0·1%).

### Ashkenazi Jewish ancestry

In AJ, pathogenic or risk variants were identified in 26·5% (620/2343) of PD individuals. Almost all genetic findings involved *GBA1* and *LRRK2*, each characterised by a predominant recurrent variant (*GBA1* p.N409S and *LRRK2* p.G2019S), alongside a smaller number of additional variants (Figures 3 and 4). Variants in *GBA1* were observed in 394 individuals (16·8%) and *LRRK2* variants in 250 individuals (10·7%); 25 of these individuals were dual *GBA1-LRRK2* variant carriers. Only one PD individual carried a *SNCA* multiplication (<0·1%).

Outside PD, variants were detected in 16·3% (16/98) of individuals with other neurological phenotypes, largely involving *GBA1*, with *LRRK2* variants identified in a small number of cases. Among controls, 10·4% (95/913) carried pathogenic and risk variants in *GBA1* and *LRRK2*.

### Latinos and Indigenous people of the Americas

Among individuals with PD, 8·7% (245/2809) carried pathogenic and risk variants across several PD-linked genes, including *GBA1* (162/2809, 5·8%), *LRRK2* (59/2809, 2·1%), *PRKN* (24/2809, 0·9%), and *SNCA* (2/2809, <0·1%); two individuals were dual *GBA1-LRRK2* carriers. Variant spectra for *GBA1* and *LRRK2* are shown in Figures 3 and 4, respectively. Of individuals with other phenotypes, two individuals carried biallelic variants in *ATP13A2* (2/26, 7·7%). In controls, pathogenic or risk variants were identified in 1·8% (30/1654), exclusively involving *GBA1*.

### Central Asian ancestry

Genetic findings in PD in CAS were identified in 8·2% (105/1284), primarily attributable to PD risk-associated variants in *GBA1* (75/1284, 5·8%) and *LRRK2* (23/1284, 1·8%) (Figures 3 and 4). Pathogenic causative variants were rare and included single individuals with variants in *LRRK2* and *RAB32* (<0·1% each, respectively), as well as *PRKN* variants identified in seven individuals (0·5%). Two individuals carried variants in more than one PD-associated gene (*PRKN*-*GBA1* and *LRRK2* risk-*GBA1*).

Genetic findings among individuals with other neurological phenotypes were rare (2/131, 1·5%) and involved one *GBA1* variant and one *LRRK2* risk variant carrier. In controls, pathogenic or risk variants were detected in 3·0% (44/1480), including *GBA1* variants (n=29), PD risk-associated *LRRK2* variants (n=14), and a single biallelic *PRKN* carrier.

### East Asian ancestry

In EAS, 17·6% (840/4773) of PD individuals carried causative or risk variants, the majority harbouring one or both of the two *LRRK2* risk variants R1628P and G2385R, identified in a total of 601 individuals (12·6%). *GBA1* variants were the next most frequent with 190 carriers (4·0%), with a broad and heterogeneous variant spectrum (Figure 3), including over 40 distinct variants. Pathogenic *LRRK2* variants were rare (n=15, 0·3%; Figure 4), and additional causative variants were observed in *SNCA* (n=4, <0·1%), *VPS35* (n=5, 0·1%), *PINK1* (n=17, 0·4%), and *PRKN* (n=17, 0·4%). Notably, all biallelic *PINK1* variant carriers harboured the same p.L347P variant in the homozygous state, while this variant was also observed in the heterozygous state in PD patients (13/4773, 0·3%), controls (6/2809, 0·2%), and unaffected family members (4/80, 5·0%). 25 individuals with PD were dual carriers, most frequently a *LRRK2* risk variant in combination with *GBA1*.

Genetic findings were also observed in 9·1% (60/662) of individuals with other neurological phenotypes, again most commonly involving *LRRK2* risk variants (n=51), with fewer *GBA1* variant carriers (n=9). Similarly, among the 6·7% (187/2809) of controls with a genetic finding, *LRRK2* risk variants were most frequent (n=170).

### European ancestry

Genetic findings were identified in 9·9% (4054/40773) of individuals with PD and across a range of different PD-associated genes. *GBA1* variants accounted for the largest proportion, with 3431 carriers (8·4%), most commonly involving the established PD risk variants p.E365K and p.T408M, but with a heterogeneous overall variant spectrum (Figure 3). Pathogenic *LRRK2* variants were the second most frequent finding and identified in 1·1% (453/40773) individuals, encompassing a broad range of variants, including several predominantly or exclusively observed in this ancestry (Figure 4). *LRRK2* risk variants were detected in a small subset (n=21, <0·1%). About 0·1% (49/40773) were dual carriers, most frequently involving *GBA1* in combination with *LRRK2*. Beyond *GBA1* and *LRRK2*, variants were detected in several additional PD-associated genes, including *SNCA* (n=31, <0·1%), *VPS35* (n=8, <0·1%), and *RAB32* (n=17, <0·1%). Among recessive genes, biallelic variants were most frequently identified in *PRKN* (126/40773, 0·3%), whereas *PINK1* (n=5) and *PARK7* (n=3) variants were rare (both <0·1%, respectively).

Genetic findings were also observed outside PD, including 6·3% (263/4153) of individuals with other neurological phenotypes and 6·2% (1337/26 761) of controls. In both groups, variants most often involved *GBA1*, with fewer pathogenic *LRRK2* or other variants.

### Finnish ancestry

We observed a genetic finding in 20·5% of FIN individuals with PD (26/127). One individual carried a biallelic *PRKN* with a co-occurring *GBA1* variant, while the remaining individuals carried *GBA1* variants only. Similarly, genetic findings were confined to *GBA1* among individuals with other neurological phenotypes (3/9, 33·3%) and controls (4/22, 18·2%).

### Middle Eastern ancestry

Genetic findings in PD among the MDE ancestry individuals were identified in 11·7% (158/1347) and distributed across multiple PD-associated genes. Most frequent were variants in *GBA1* (n=73, 5·4%) and *LRRK2* (n=59, 4·4%), with the *LRRK2* variant spectrum largely driven by p.G2019S (Figure 4), whereas *GBA1* showed a more heterogeneous spectrum (Figure 3). Additional findings included variants in *PRKN* (n=17, 1·3%), *PINK1* (n=11, 0·8%), *RAB32* (n=4, 0·3%), and *SNCA* (n=1, <0·1%). Seven individuals were dual carriers of *GBA1* and *LRRK2* variants.

Genetic findings in individuals with other phenotypes included *GBA1* in two cases and *LRRK2* in one (overall 3/28, 10·7%). Among controls, pathogenic or risk variants were detected in 1·9% (25/1340), most frequently involving *GBA1*.

### South Asian ancestry

Pathogenic or risk variants were identified in 6·1% (49/807) of PD patients. *GBA1* variants were the most frequent findings (n=39, 4·8%), with p.L483R being the most frequent variant (Figure 3). Variants in recessive PD genes were also observed, including *PRKN* (n=5, 0·6%) and *PINK1* (n=4, 0·5%), while pathogenic variants in *LRRK2* and *VPS35* were each identified in a single individual only (0·1% each, respectively).

Among individuals with other phenotypes (1/74; 1·4%) and controls (4/440; 0·9%), only *GBA1* variants were identified.

### Single heterozygous variants in recessively inherited genes

Across all ancestries, we identified 1182 individuals with PD carrying single heterozygous pathogenic variants in recessive genes (1182/58559, 2·0%), most frequently in *PRKN* (n=1064), followed by *PINK1* (n=40), *ATP13A2* (n=22), and *FBXO7* (n=21) whereas variants in other genes (*JAM2*, *PARK7*, *SYNJ1*, and *VPS13C*) were rare. A substantial proportion of these, 41·7%, had an AAO ≤50 years (377/904; AAO missing for n=278). The overall proportion of individuals with an AAO ≤50 years among the entire PD group included in this study was 26·4% (11814/44768; AAO missing for n=13791). In comparison, the frequency of single heterozygous pathogenic variants in recessive genes in controls was 1·4% (583/41239). Eleven individuals carried two heterozygous *PRKN* variants but had an AAO >60 years or the AAO was unavailable, so we did not interpret them as likely compound heterozygous without further confirmation.

## Discussion

We performed a comprehensive analysis of causative and risk variants in PD-associated genes across diverse populations. Our work represents the largest ancestry-informed assessments to date, including ∼30% of non-European and non-Ashkenzi Jewish ancestry individuals, offering insights into the population-specific genetic architecture of PD. This is important as multiple ongoing or planned clinical trials target proteins encoded by PD-linked genes.^12,13^ Ensuring these advances are inclusive requires a detailed understanding of genetic variation beyond European ancestry.^14^

The overall yield of causal variants across PD individuals was ∼2%, with substantial variations across ancestries ranging from about 0·5% in AAC and AFR to >10% in AJ. Including risk variants raised the yield to ∼14%, aligning with prior studies predominantly based on European individuals.^15,16^ This increase was largely driven by *GBA1*, except in EAS, where the *LRRK2* risk variants p.R1628P and p.G2385R dominated. Notably, yields were substantially lower among individuals with other neurodegenerative or movement disorders, likely due to the marked heterogeneity of this group, encompassing multiple phenotypes with distinct underlying genetic etiologies beyond the genes assessed in this study. Interestingly, the proportion of individuals carrying causal or risk variants in controls exceeded that observed in other phenotypes across most ancestries. This pattern may partly reflect the substantially smaller and uneven sample sizes of these other phenotypic groups.

*GBA1* risk variants were most frequent overall and identified across all populations, albeit with distinct ancestry-specific variant spectra: p.N409S was the most common variant in AJ, whereas p.E365K and p.T408M were more frequent in EUR, FIN, and CAS, but rare in AFR, AAC, and EAS. The variant spectrum in EAS was broad, but p.L483P appeared more frequent than in other populations. The intronic variant rs3115534-G was by far the most frequently observed in AAC and AFR, as previously reported.^17^ This variant represents a clear example of an ancestry-specific *GBA1* risk allele that would have remained undetected in studies restricted to European-ancestry populations, underscoring the necessity of ancestry-diverse genetic investigations. Collectively, these findings emphasize the importance of *GBA1* as a globally relevant therapeutic target, an important observation given that most trials targeting glucocerebrosidase are conducted in countries with a high representation of European or Ashkenazi Jewish populations.^4^

*LRRK2* variants also showed ancestral variability. p.G2019S was detected across multiple ancestries, with the highest frequencies in MDE (partially reflecting North African Berbers), AMR, and AJ.^18,19^ While p.G2019S was also the most frequent *LRRK2* variant in EUR, the mutational spectrum was notably broader and included p.R1441C, p.R1441G, and p.L1795F with suggested founder effects in European sub-populations.^20–23^ In contrast, p.G2019S was largely absent from Asian populations, where the risk variants p.R1628P and p.G2385R were most frequent,^24^ especially in EAS. The most frequent pathogenic *LRRK2* variant identified in the EAS population was p.R1067Q, only recently proven to be disease-relevant and shown to be enriched in East Asians.^25^ Together, these observations illustrate that, while *LRRK2* is a globally relevant therapeutic target, ancestry-specific studies are essential to capture distinct variant spectra, including the possibility of yet-unidentified *LRRK2* variants in under-studied African populations. *LRRK2* variants differ in their impact on kinase activity,^26^ which may inform clinical trial design and therapeutic targeting. Investigating these actionable variants globally is key, particularly given the ongoing LRRK2-targeted trials.

*SNCA* variants were overall rare but identified across multiple ancestries, in line with previous reports.^27^ While *SNCA* multiplications were identified in several populations, missense variants (p.A53T and p.G51D) were predominantly observed in EUR. Other known autosomal dominant forms of PD caused by *VPS35* p.D620N or *RAB32* p.S71R were rare. *RAB32* variants were only detected in EUR and MDE individuals, more frequent in the latter, while *VPS35* variant carriers were observed across three different populations, with a relatively slightly higher proportion in EAS and SAS compared to EUR. Given the low sample size of some populations, especially with sequencing data, these findings need cautious interpretation. Nonetheless, detecting *VPS35* and *RAB32* variants across multiple ancestries underscores the importance of continued investigation in larger, diverse cohorts.

Investigating recessive forms of PD across populations, we found *PRKN* to be the most frequently implicated gene, followed by *PINK1*, while only three European-ancestry *DJ-1/PARK7* carriers were observed. Biallelic *PRKN* variants were identified across nearly all populations, except AJ. In Asian populations (EAS, CAS, and SAS), *PRKN* variants were more frequent than pathogenic *LRRK2* variants, although this warrants cautious interpretation given the small sample sizes in some ancestries. Identifying *PRKN* carriers across multiple ancestries reinforces its global relevance, as preclinical efforts increasingly explore therapeutic strategies targeting Parkin (encoded by *PRKN*) or its associated pathways.^12^ While *PINK1* variants were rare, we identified a notable proportion of MDE, EAS, and SAS carriers. All EAS *PINK1* carriers harbored p.L347P, a variant known to be enriched in this population.^28^ We further identified numerous individuals carrying single heterozygous pathogenic variants in recessive PD genes. While these are not causative, many carriers had an early AAO ≤50 years, suggesting a second pathogenic variant, particularly a complex structural variant not captured by genotyping or short-read sequencing.

This global analysis underscores the clinical and translational importance of extending PD genetics research beyond European ancestry. Ancestry-informed genetic studies improve diagnostic accuracy, variant interpretation, and genotype-phenotype correlations, with direct implications for prognosis and genetic counselling. Our findings further show that variants predominantly identified in European populations often contribute only modestly in other ancestries, suggesting additional ancestry-specific variants and highlighting that our current understanding of the genetic architecture of PD remains incomplete. From a translational perspective, such diversity is essential for equitable clinical trial design, accurate identification of trial-eligible individuals, and the global applicability of emerging genetically targeted therapies. Together, these data provide a robust reference framework and a foundation for future in-depth ancestry-specific analyses and precision-medicine efforts in PD.

### Limitations

Our study focused on variants in known PD genes, primarily identified in European populations. This and the predominance of European-ancestry data in resources like ClinVar may bias variant interpretation in non-European groups. While our study allowed for a large-scale, clinically relevant assessment of those known variants, it limits the discovery of novel PD genes and variants that may be more relevant in other populations. Systematically studying large, well-powered non-European ancestry cohorts using sequencing data will be important to capture the full global spectrum of novel variation. Another important area will be investigating variants of uncertain significance in known PD genes. Some variants may be pathogenic in specific populations but are currently classified as of uncertain significance due to limited ancestry-specific reference data and low statistical power. Ancestry-aware (re)classification using variant frequency, functional, and segregation data will be key for establishing pathogenicity, improving genetic counseling and enabling trial access.^23,25^

While our study is large and ancestrally diverse, some populations were of limited sample size. This may inflate the relative contribution of known genes in well-powered populations, and is particularly relevant for groups with limited WGS data, since select variants are not captured by genotyping (e.g., *RAB32* p.S71R and select *GBA1* variants).

The detection of compound-heterozygous variants was limited by the absence of phased data. We conservatively considered carriers of two heterozygous pathogenic variants in recessive genes with an AAO ≤60 years as likely compound heterozygous, but validation is needed. Notably, although genetically inferred ancestry provides a more robust framework than geographic labels, the broad ancestry categories still encompass substantial heterogeneity and may reflect cohort-specific recruitment patterns. Nonetheless, we believe this approach represents the most transparent and scalable strategy currently feasible for large, harmonised cross-ancestry analyses.

Lastly, findings were generated in a research setting; while we employed quality control measures and WGS validation where possible, further confirmation is needed.

## Conclusion

In conclusion, this study represents the largest global assessment of the genetic spectrum of PD, offering crucial insights into the population-specific genetic architecture of PD and underscoring the critical need to expand PD genetics research beyond European ancestry. Ancestry-informed analyses enhance diagnostic precision and risk interpretation and are essential to ensure equitable access to genetically stratified clinical trials and emerging precision therapies. Addressing current gaps in data diversity and global genetic representation is not optional but fundamental to translating genetic discovery into effective, inclusive precision neurology on a global scale.

## Author contributions

LML, ZHF, and NK accessed and verified the data reported in this manuscript. LML, SoB, HH, RK, KRK, SYL, NEM, WMYM, AJN, RO, NUO, SUR, CS, SS, AHT, ZT, BT, EMV, NZ, HRM, and CK carried out study assessments for participants, including clinical data and biospecimen collection. ZHF, MBM, MJK, HLL, KSL, MAN, and DV were responsible for processing, performing quality control, and harmonizing genotyping and sequencing data from the Global Parkinson’s Genetics Program. HI and LJ were responsible for processing and harmonizing clinical data from the Global Parkinson’s Genetics Program. KAB, ShB, MLD, SJ, JJ, EJS, and MT contributed to the acquisition of data. MBM, HLL, KL, SBC, CB, and AS contributed to the analysis of genetic data. LML and CK conceptualised the study. PH, LS, and JT supported the execution of the study. CB, AS, HRM, and CK supervised the study. LML drafted the manuscript. All authors had full access to all the data in the study and had final responsibility for the decision to submit the manuscript for publication.

## Data and Code Availability

Data used in the preparation of this article were obtained from the Global Parkinson’s Genetics Program (GP2; https://gp2.org). Specifically, we used Tier 2 data from GP2 release 11 (https://doi.org/10.5281/zenodo.17753486). Tier 1 data can be accessed by completing a form on the Accelerating Medicines Partnership in Parkinson’s Disease (AMP^®^-PD) website (https://amp-pd.org/register-for-amp-pd). Tier 2 data access requires approval and a Data Use Agreement signed by your institution. Qualified researchers are encouraged to apply for direct access to the data through AMP PD.

All code generated for this article, and the identifiers for all software programs and packages used, are available on GitHub (https://github.com/GP2code/GP2-global-genetic-variant-landscape) and were given a persistent identifier via Zenodo (DOI 10.5281/zenodo.15699539).

A detailed list of all identified variant carriers, including corresponding anonymised GP2-IDs, variant details as well as basic demographic and clinical characteristics are available to qualified researchers and upon reasonable request from the corresponding author.

## Data Availability

Data used in the preparation of this article were obtained from the Global Parkinson's Genetics Program (GP2; https://gp2.org). Specifically, we used Tier 2 data from GP2 release 11 (https://doi.org/10.5281/zenodo.17753486). Tier 1 data can be accessed by completing a form on the Accelerating Medicines Partnership in Parkinson's Disease (AMP-PD) website (https://amp-pd.org/register-for-amp-pd). Tier 2 data access requires approval and a Data Use Agreement signed by your institution. Qualified researchers are encouraged to apply for direct access to the data through AMP PD.

## Acknowledgments

This project was supported by the Global Parkinson’s Genetics Program (GP2; https://gp2.org). GP2 is funded by the Aligning Science Across Parkinson’s (ASAP) initiative and implemented by The Michael J. Fox Foundation for Parkinson’s Research (MJFF). For a complete list of GP2 members see doi.org/10.5281/zenodo.7904831.

This research was supported in part by the Intramural Research Program of the National Institutes of Health (NIH). The contributions of the NIH authors are considered Works of the United States Government. The findings and conclusions presented in this paper are those of the authors and do not necessarily reflect the views of the NIH or the U.S. Department of Health and Human Services.

We thank Yuliia Kanana for her valuable support in processing and handling the samples used in this study. We are grateful for the important support from all study participants and their families as well as for contributions of all collaborators, brainbank and biobanks.

