## Supplementary Material for "The Global Landscape of Genetic Variation in Parkinson’s disease: Multi-Ancestry Insights into Established Disease Genes and their Translational Relevance"

[#] Shared last authors

**Correspondence to**
Lara M. Lange
Laboratory of Neurogenetics
National Institute on Aging (NIA), National Institutes of Health (NIH)
9000 Rockville Pike
Bethesda, MD 20892,
United States of America


or Christine Klein
Institute of Neurogenetics
University of Luebeck
Ratzeburger Allee 160
23538 Luebeck,
Germany


#### Table of Content

| | Page |
|---|---|
| **Supplementary Methods** | |
| Ethics declaration | 4 |
| Whole-genome sequencing (WGS) data (GP2 and AMP-PD) processing | 4 |
| Clinical exome sequencing data (PDGENEration) processing | 4 |
| Genome-wide genotyping data from the NeuroBooster Array (NBA) processing | 4 |
| Variants of interest and pathogenicity evaluation | 4-5 |
| Validation of genetic findings and concordance check | 5 |
| Additional statistical analyses | 5 |
| **Supplementary References** | 5-6 |
| **Supplementary Figures** | |
| Supp Figure 1. Ancestry-specific distribution of genetic findings in Parkinson's disease and controls. | 7-8 |
| Supp. Figure 2: Ancestry-Stratified Age at Onset in Idiopathic, *GBA1*-, and *LRRK2*-Associated Parkinson's Disease. | 9 |
| Supp. Figure 3: Cluster plots. | 10-20 |
| **Supplementary Tables** | |
| Supp. Table 1: Overview of included samples investigated in this study. | 21-22 |
| Supp. Table 2: Summary of genetic findings across all ancestries. | 23-24 |
| Supp. Table 3: Summary of genetic findings across all ancestries in atypical parkinsonism genes. | |
| Supp. Table 4: Results of linear regression analysis comparing age at onset (AAO) between idiopathic PD (IPD, reference group) and *LRRK2*- and *GBA1*-associated PD across ancestries. | 25<br>26 |

#### Supplementary Methods

##### Ethics declaration

This study was approved by ethics committees or institutional review boards of all participating sites and conducted in accordance with their ethical standards. Informed consent for study participation was obtained from all participants.

##### Whole-genome sequencing (WGS) data (GP2 and AMP-PD) processing

We used WGS data generated as part of GP2 Data Release 11 (https://doi.org/10.5281/zenodo.17753486). All samples were genome sequenced to an average of 30x coverage with 150bp paired-end reads following Illumina's TruSeq PCR-free library preparation protocol. We followed AMP-PD's functional equivalence pipeline[1] to produce the sequence alignment against the GRCh38DH reference genome. DeepVariant v.1.6.1[2] (https://github.com/google/deepvariant) was used to generate the single-sample variant calls, and joint-genotyping was performed using GLnexus v1.4.3 (https://github.com/dnanexus-rnd/GLnexus) with the preset DeepVariant WGS configuration[3]. Genotypes were set to be missing after variant quality control defined as genotype quality >=10, read depth >=10, and heterozygous allele balance between 0.2 and 0.8, and retained high-quality variants with a call rate > 0.95 after quality control. Genetic ancestry was determined using GenoTools v1.2.3 (https://github.com/GP2code/GenoTools) using default settings[4].
Variants in the genes of interest were extracted using PLINK[5,6] and annotated with ANNOVAR[7,8]. We used the Gauchian pipeline (https://github.com/Illumina/Gauchian)[9] for WGS data as a variant caller for *GBA1*.

##### Clinical exome sequencing data (PDGENEration) processing

We used clinical exome sequencing data from 10,454 individuals with PD generated by PDGENEration[10] and released as part of GP2 Data Release 11 (https://doi.org/10.5281/zenodo.17753486). Data processing followed the same pipeline we used for WGS data (described above). We performed joint-genotyping using GLnexus v1.4.3 with the preset DeepVariant whole exome sequencing (WES) configuration and followed the same criteria for sample and variant quality used for WGS data.
Variants in the genes of interest were extracted using PLINK[5,6] and annotated with ANNOVAR[7,8].

##### Genome-wide genotyping data from the NeuroBooster Array (NBA) processing

We used raw genotyping data generated as part of GP2 Data Release 11 (https://doi.org/10.5281/zenodo.17753486). Genotyping was performed using the NeuroBooster Array (NBA; v.1.0, Illumina, San Diego, CA)[12]. Raw genotyping data underwent quality control and genetic ancestry prediction using GenoTools v1.2.3 with the default settings[4].
Variants in the genes of interest were extracted using PLINK[5,6] and annotated with ANNOVAR[7,8]. Copy number variation (CNV) analyses were performed for all samples with genotyping data as described before[13] and included screening for *SNCA* multiplications and *PRKN* deletions and duplications.

Further, we imputed rs3115534-G using the Michigan Imputation Server 2 (https://imputationserver.sph.umich.edu/)[14] by uploading chromosome 1 of each ancestry separately (GP2 release 11; https://doi.org/10.5281/zenodo.17753486). All ancestries were imputed to the 1000G Phase 3 30x (GRCh38/hg38) panel. Only ancestries with a reliable estimated squared correlation between the true allele dosage and the imputed allele dosage ($R^2$) were considered, thus, only variant carriers of African Admixed (AAC), African (AFR), Latinos and Indigenous People of the Americas (AMR), Complex Admixture (CAH), and Middle Eastern (MDE) ancestry were extracted.

##### Variants of interest and pathogenicity evaluation

We focused our analyses on variants predicted as pathogenic/likely pathogenic according to ClinVar (https://www.ncbi.nlm.nih.gov/clinvar/)[15] and/or the consensus recommendations of the American College of Medical Genetics and Genomics (ACMG)[16]. For variants with conflicting ClinVar predictions or those absent from ClinVar, pathogenicity was evaluated using Franklin (https://franklin.genoox.com) and Varsome (https://varsome.com/)[17], both based on the ACMG criteria. We added *RAB32* variant (chr6:146544084:C:G, p.S71R), that is not yet included in ClinVar and still predicted to be a variant of uncertain significance despite convincing evidence of a causal role from the literature.[18–21] We further included more common *LRRK2* variants classified as PD risk-associated based on association studies, including rs33949390 (chr12:40320043:G:C, p.R1628P) and rs34778348 (chr12:40363526:G:A, p.G2385R), given their translational relevance. Individuals harboring two pathogenic/likely pathogenic variants in recessive genes with an age at onset (AAO) ≤50 years were considered likely compound heterozygous although the phase could not be determined. In addition to causal genes, we also investigated *GBA1* variants, given their substantial translational impact. For the purpose of this study and in the context of PD, all *GBA1* variants mentioned in the main text are referred to as risk variants (including those predicted to be causal for Gaucher's disease); including variants of different severities (i.e., severe, mild, and risk). However, the different variant severities were evaluated with the *GBA1*-PD browser (https://pdgenetics.shinyapps.io/gba1browser/)[22] and their distribution is provided in Figure 2.

**Validation of genetic findings**
Performing wet-lab or CLIA-certified validation of genetic findings was out of the scope of this study. We performed a concordance check, using WGS and CES data as a validation of genotyping findings for samples that underwent both analyses, especially for rare variants. Further, we generated cluster plots to evaluate and ensure adequate genotyping performance of identified variants (Supplementary Figure 2). Only variants with sufficient validation or reliable cluster plots were included in our analyses.

**Additional statistical analyses**
We performed a linear regression to compare the ages at onset (AAO) between individuals with idiopathic PD and individuals with either *LRRK2*-linked PD or *GBA1*-associated PD. We considered all individuals that did not carry a causative of high-risk variant in the genes of interest investigated in this study as "idiopathic". AAO was analysed using ancestry-stratified multivariable linear regression models (ordinary least squares). AAO was modelled as a continuous outcome with disease group as a categorical predictor, using IPD as the reference group, and adjusted for sex. Regression coefficients represent mean differences in AAO relative to IPD within each ancestry. A p-value of <0.05 was considered statistically significant.

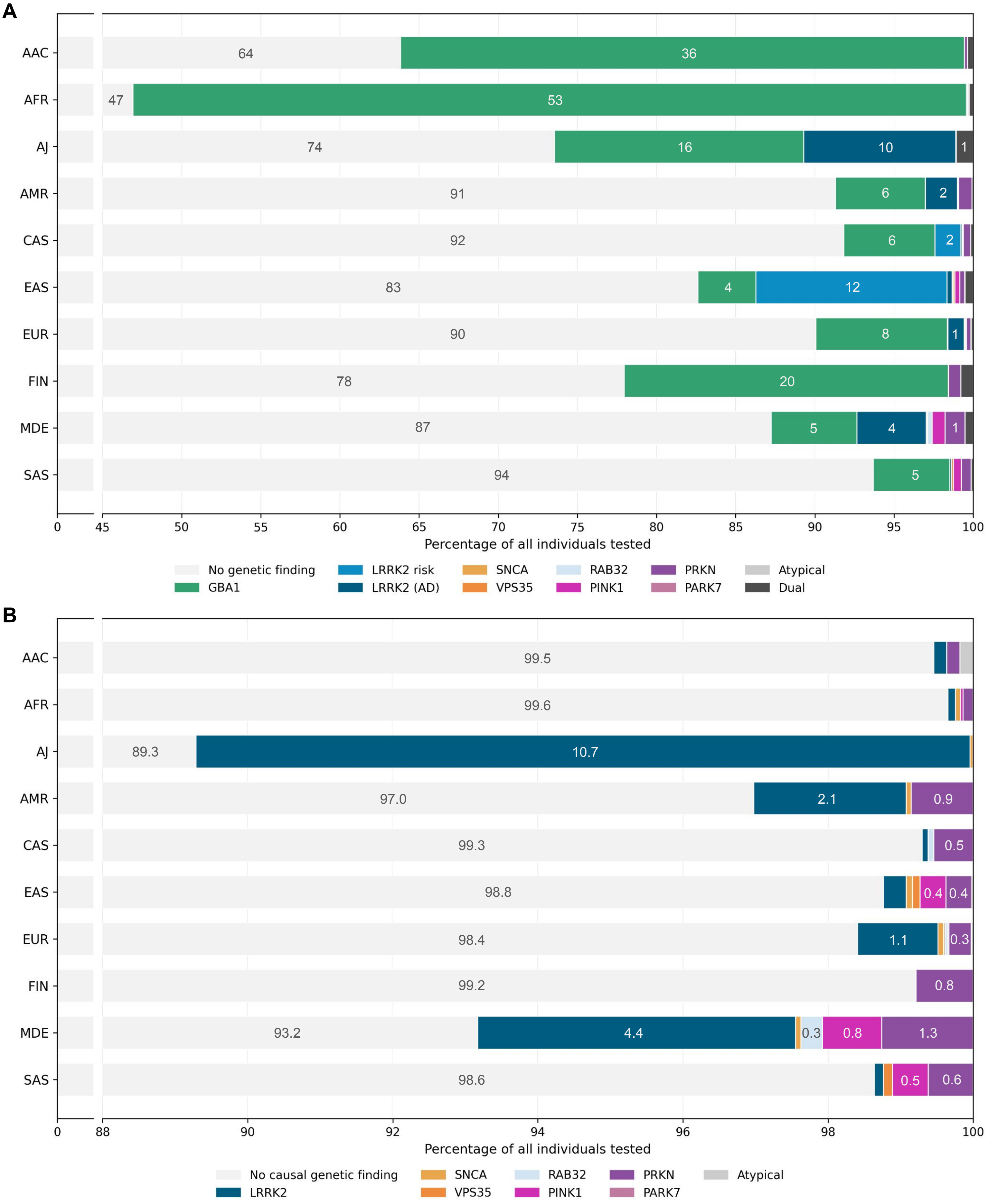
A
AAC
AFR
AJ
AMR
CAS
EAS
EUR
FIN
MDE
SAS
64
36
47
53
74
16
10
1
91
6
2
92
6
2
83
4
12
90
8
1
78
20
87
5
4
1
94
5
0
45
50
55
60
65
70
75
80
85
90
95
100
Percentage of all individuals tested
No genetic finding
GBA1
LRRK2 risk
LRRK2 (AD)
SNCA
VPS35
RAB32
PINK1
PRKN
PARK7
Atypical
Dual
B
AAC
AFR
AJ
AMR
CAS
EAS
EUR
FIN
MDE
SAS
99.5
99.6
89.3
10.7
97.0
2.1
0.9
99.3
0.5
98.8
0.4
0.4
98.4
1.1
0.3
99.2
0.8
93.2
4.4
0.3
0.8
1.3
98.6
0.5
0.6
0
88
90
92
94
96
98
100
Percentage of all individuals tested
No causal genetic finding
LRRK2
SNCA
VPS35
RAB32
PINK1
PRKN
PARK7
Atypical

C

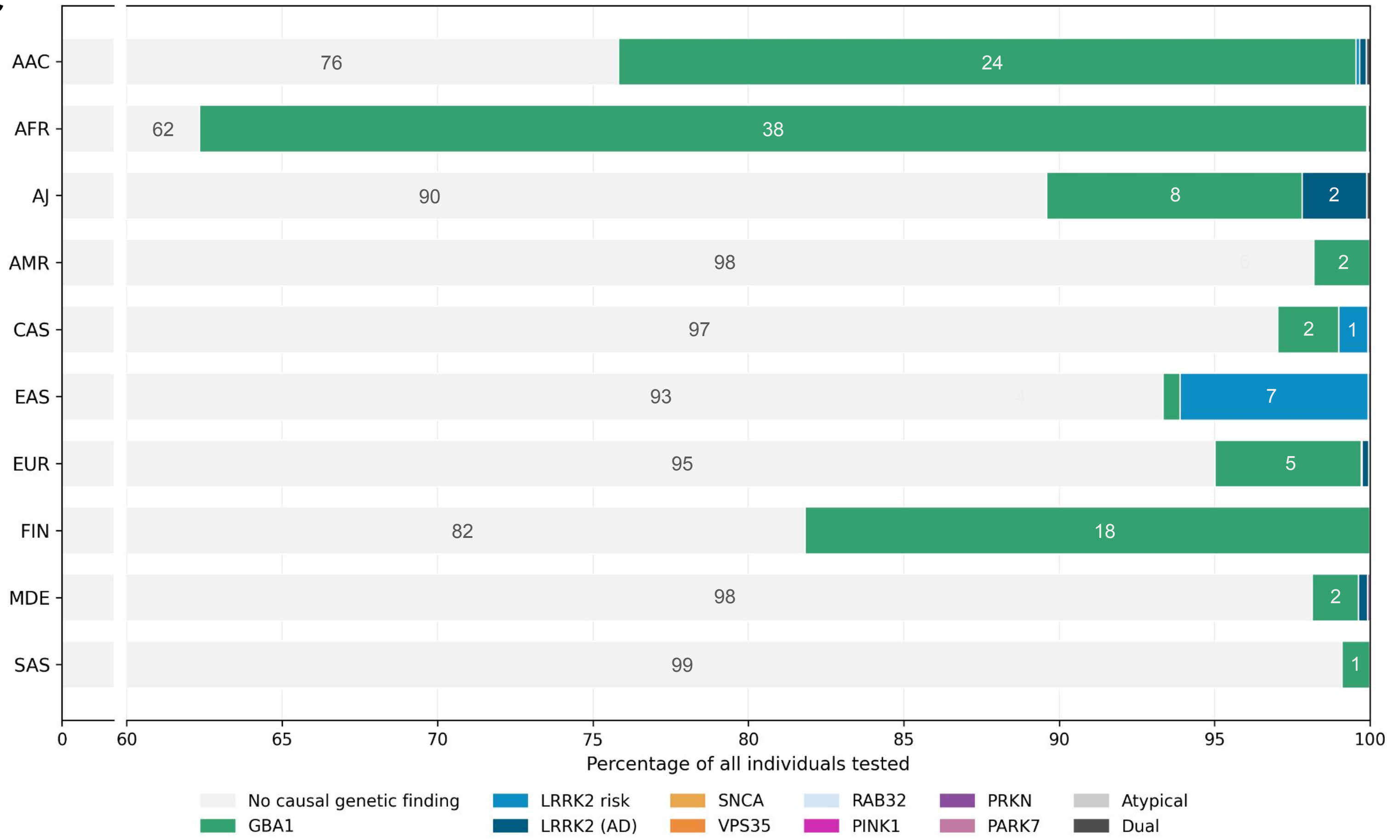

D

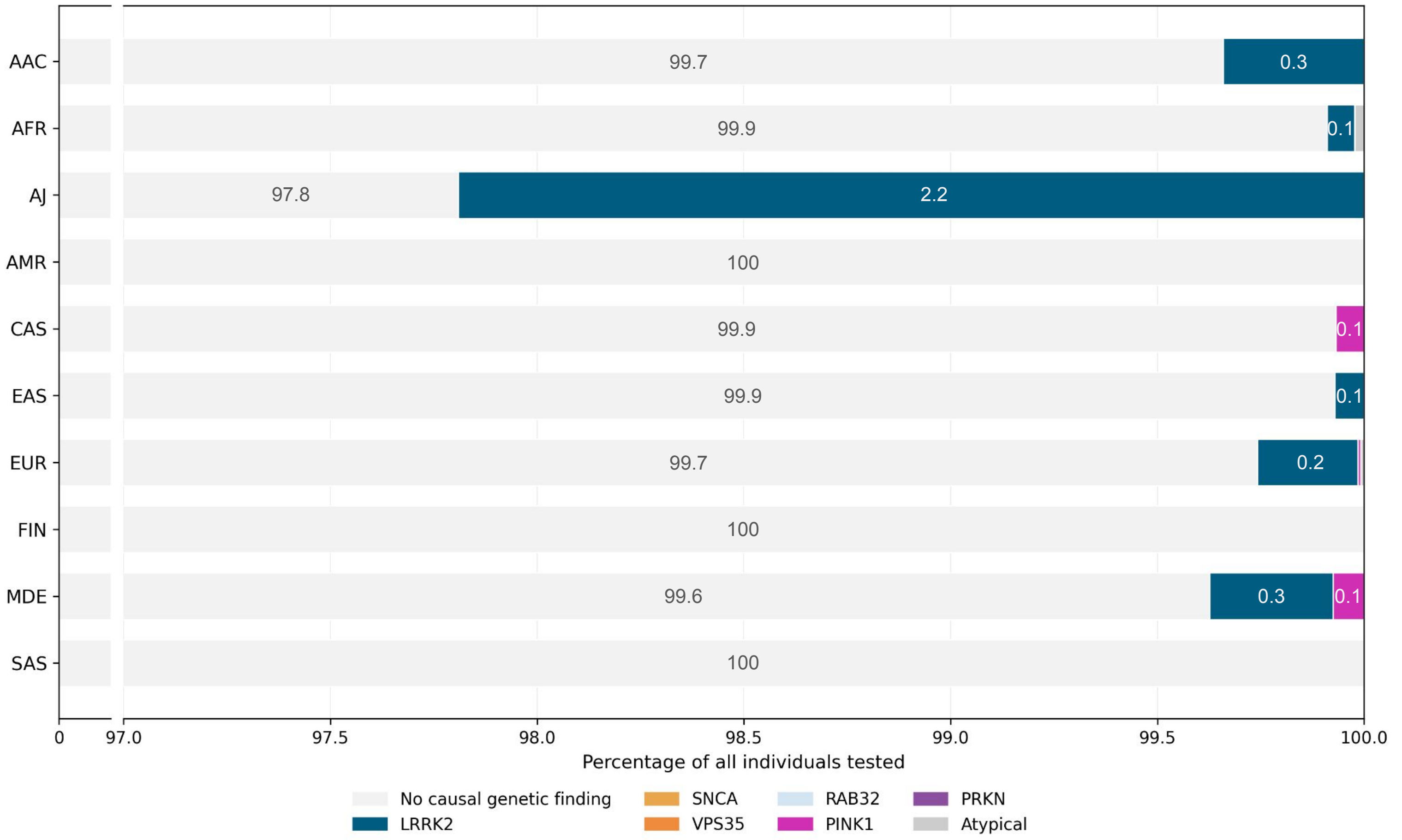

**Supplementary Figure 1. Ancestry-specific distribution of genetic findings in Parkinson's disease and controls.** Horizontal stacked bar charts displaying the proportion of individuals with a genetic finding out of all tested individuals in each ancestry group. Panels display (A) Parkinson's disease (PD) including risk-associated variants, (B) PD excluding risk-associated variants, (C) controls including risk-associated variants, and (D) controls excluding risk-associated variants. To improve visualisation of low-frequency categories, x-axes are truncated with an axis break, with expanded scaling applied to values beyond the cut-off. Numbers inside bars indicate percentages. AAC = African Admixed, AFR = African, AJ = Ashkenazi Jewish, AMR = Latinos and Indigenous people of the Americas, CAH = Complex Admixture, CAS = Central Asian, EAS = East Asian, EUR = European, FIN = Finnish, MDE = Middle Eastern, SAS = South Asian

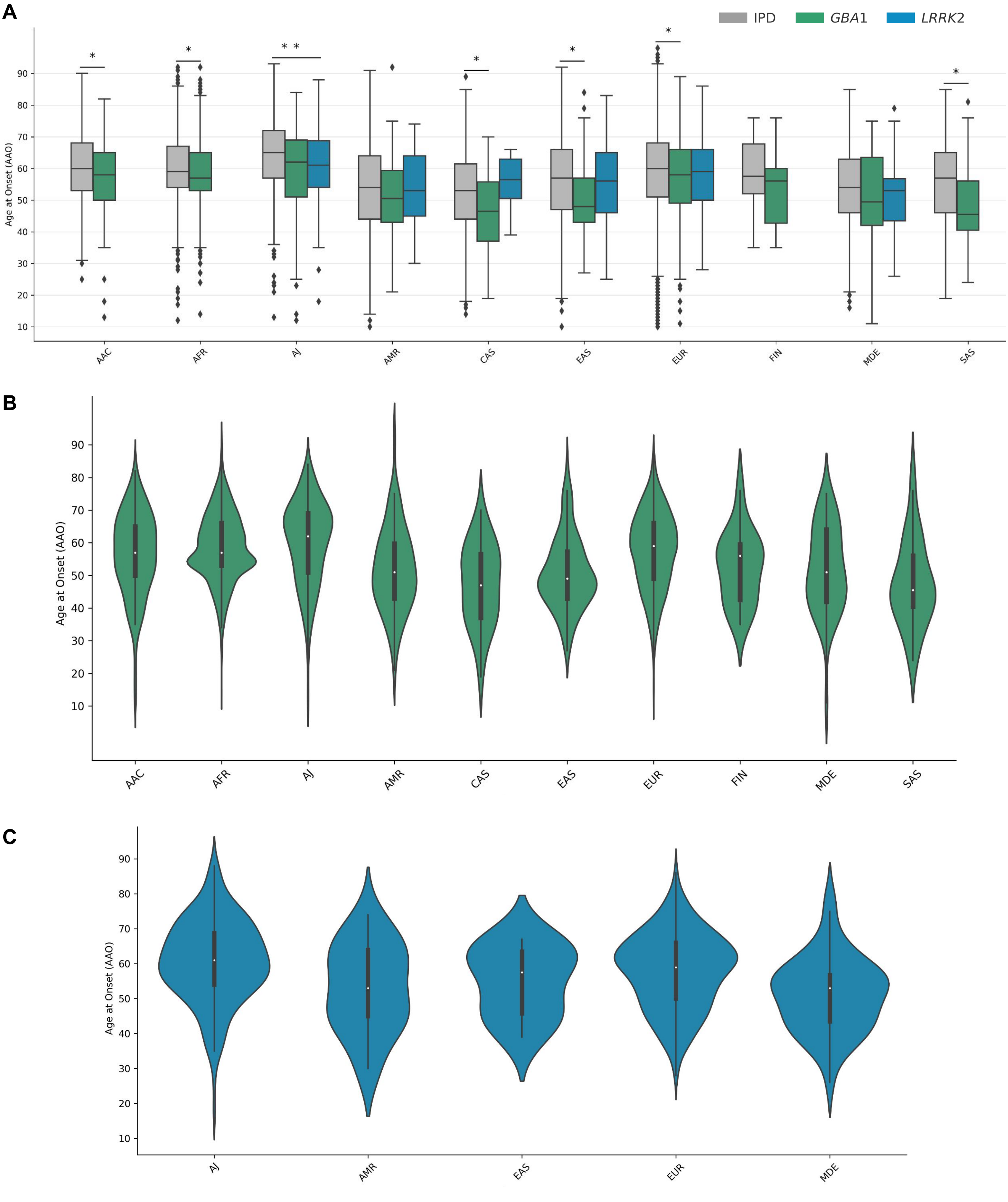

**Supplementary Figure 2. Ancestry-Stratified Age at Onset in Idiopathic, *GBA1*-, and *LRRK2*-Associated Parkinson's Disease.** Panel A shows boxplots of ages at onset (AAO) for individuals with idiopathic Parkinson's disease (IPD; grey), *GBA1*-associated PD (green), and *LRRK2*-linked PD (blue) grouped by ancestry. Boxes indicate the interquartile range (IQR), horizontal lines represent the median, and whiskers show the range excluding outliers. Outliers are plotted as individual points. Statistically significant group differences within ancestry based on linear regression (adjusted for sex) are indicated by asterisks. Panels B and C display violin plots of AAO for *GBA1*-associated PD (Panel B; green) and *LRRK2*-linked PD (Panel C; blue) across ancestries. Each violin reflects the distribution and median AAO within each ancestry group. Only ancestries with sufficient available data are shown.

AAC = African Admixed, AFR = African, AJ = Ashkenazi Jewish, AMR = Latinos and Indigenous people of the Americas, CAS = Central Asian, EAS = East Asian, EUR = European, FIN = Finnish, MDE = Middle Eastern, SAS = South Asian

### Supplementary Figure 3

### Cluster plots *PINK1* variants

##### *PINK1* p.Q129* (excluded)

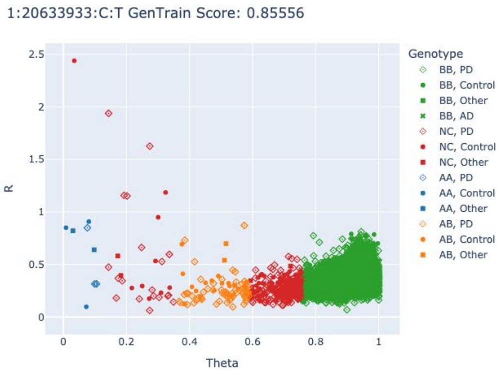

##### *PINK1* p.V170G (excluded)

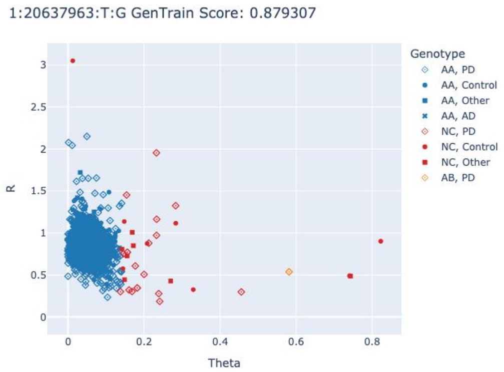

##### *PINK1* p.G309D (excluded)

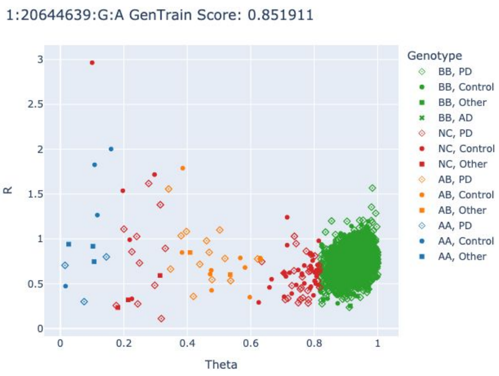

##### *PINK1* p.G309D (excluded)

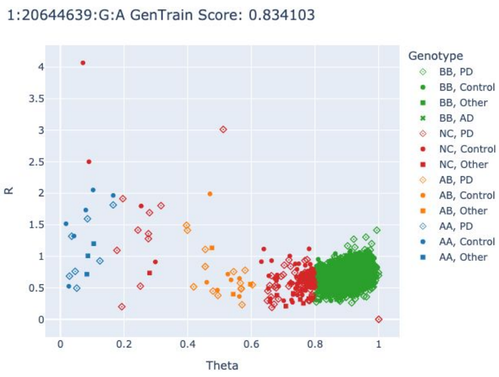

##### *PINK1* p.L347P

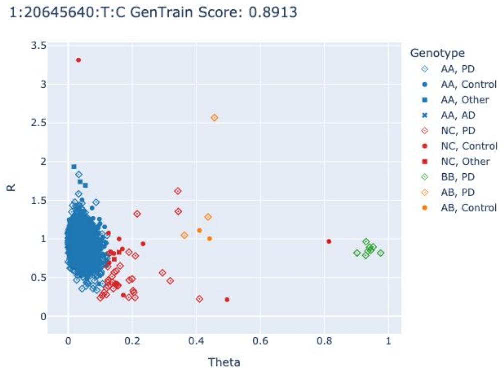

##### *PINK1* p.L347P

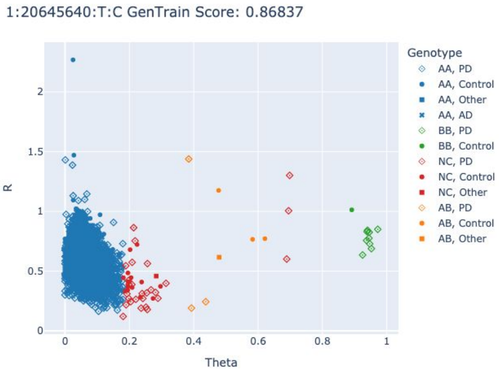

##### *PINK1* p.L347P

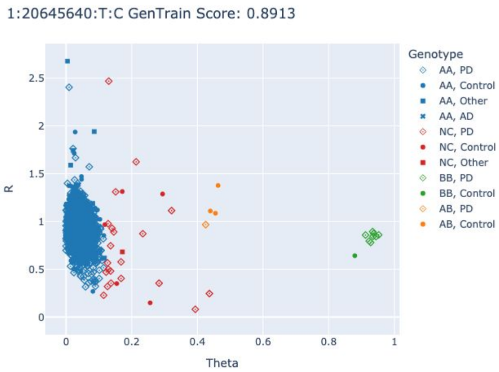

##### *PINK1* p.Q456*

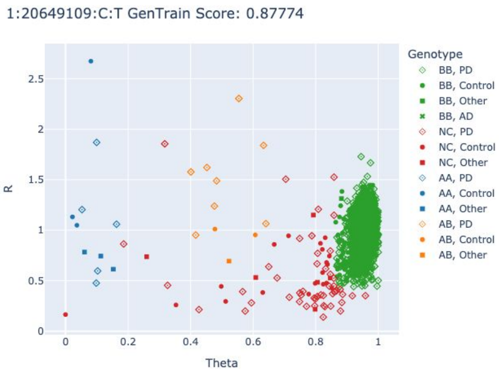

##### *PINK1* p.Q456*

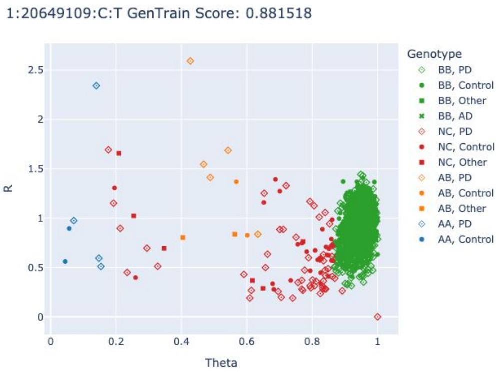

### Cluster plots *GBA1* variants

#### *GBA1* p.R535H

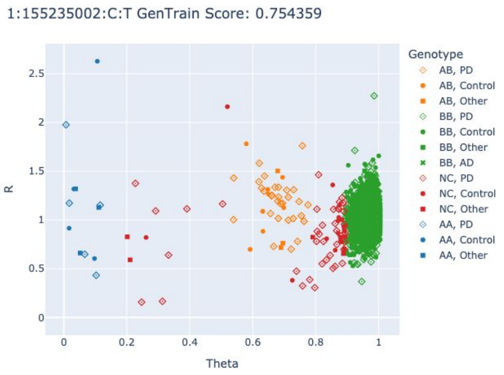

#### *GBA1* p.R502H

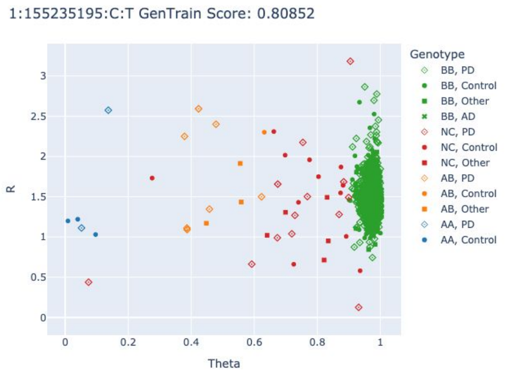

#### *GBA1* p.R502H

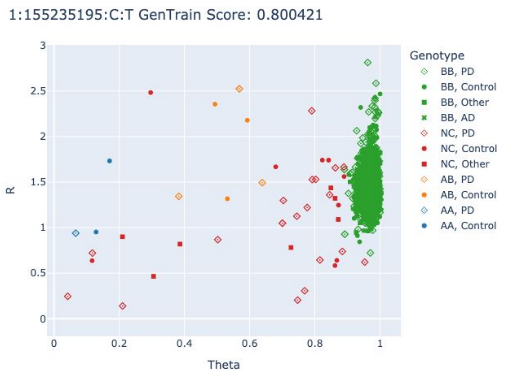

#### *GBA1* p.R502H

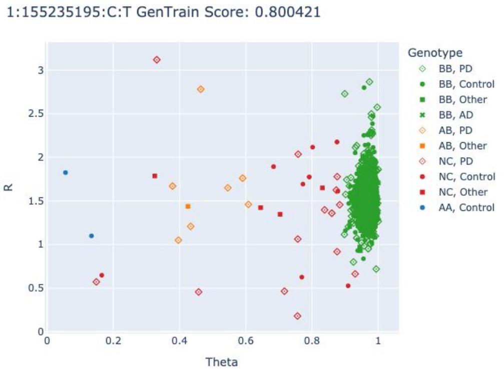

#### *GBA1* p.R502C

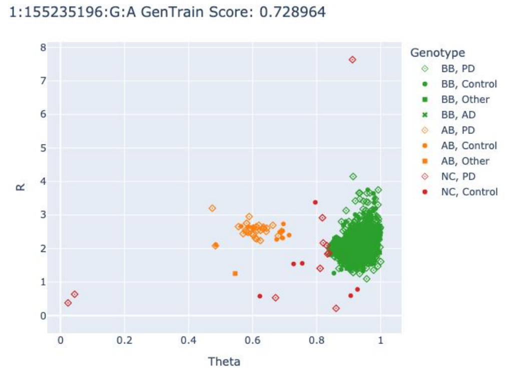

#### *GBA1* p.R502C

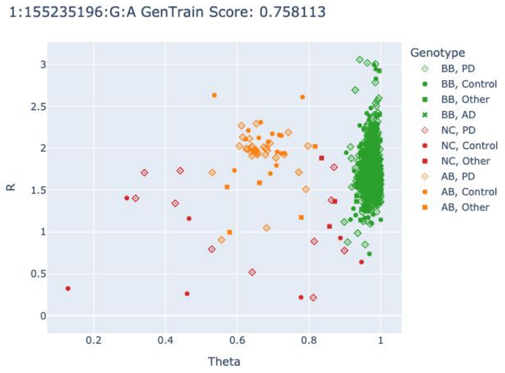

#### *GBA1* p.V499L

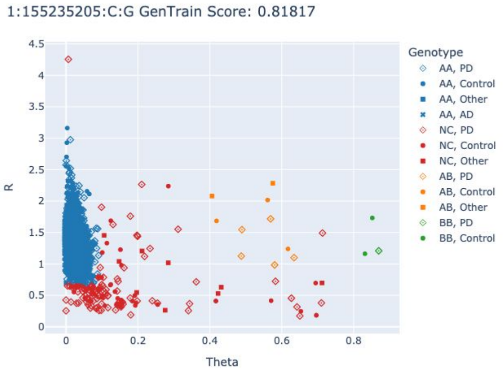

#### *GBA1* p.V499M

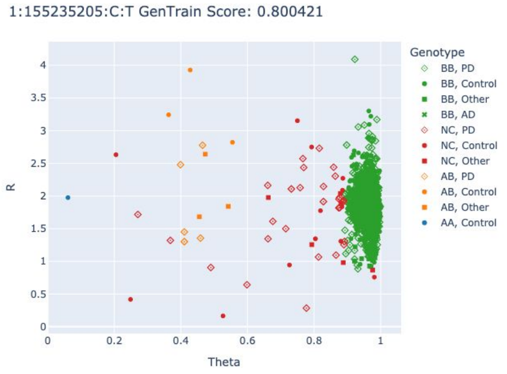

#### *GBA1* p.V499M

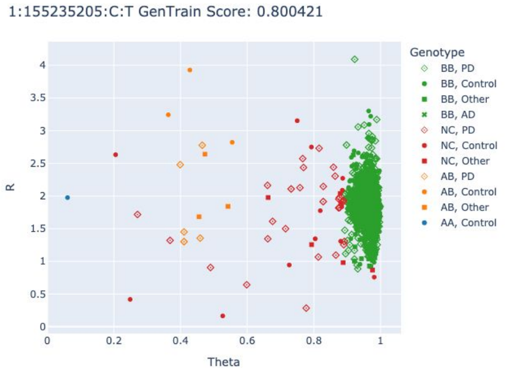

#### *GBA1* p.D482N

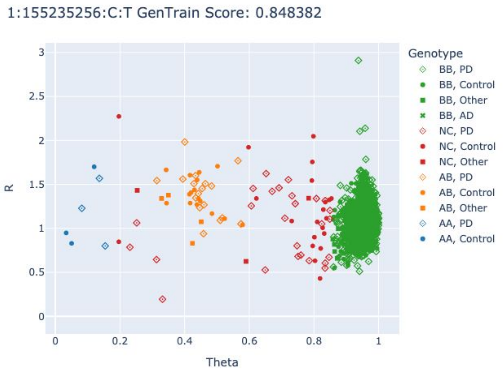

#### *GBA1* p.D482N

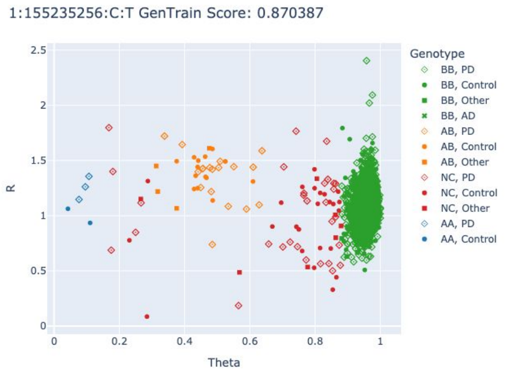

#### *GBA1* p.V433L

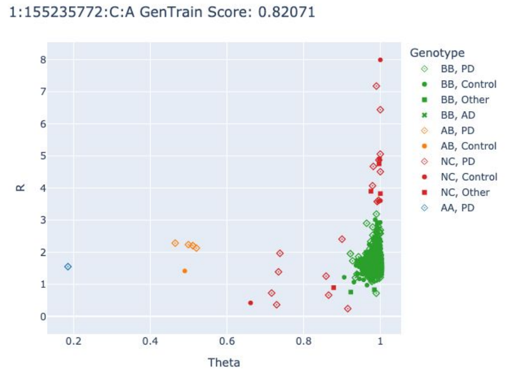

#### *GBA1* p.V433L

1:155235772:C:A GenTrain Score: 0.767985

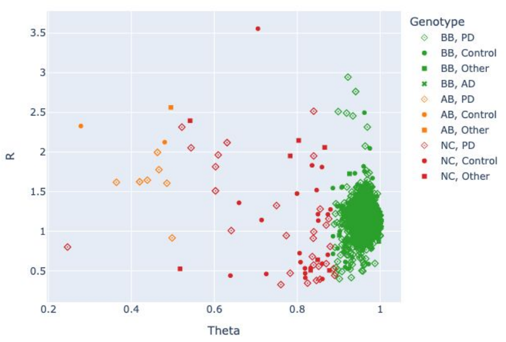

#### *GBA1* p.V433L

1:155235772:C:A GenTrain Score: 0.762123

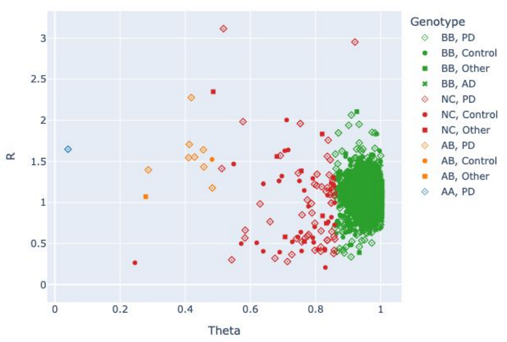

#### *GBA1* p.W432*

1:155235773:C:T GenTrain Score: 0.808818

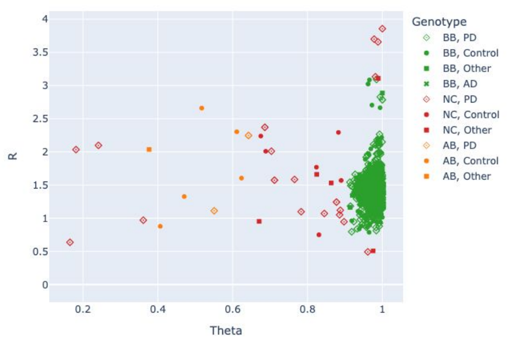

#### *GBA1* p.N431S (excluded)

1:155235777:T:C GenTrain Score: 0.800421

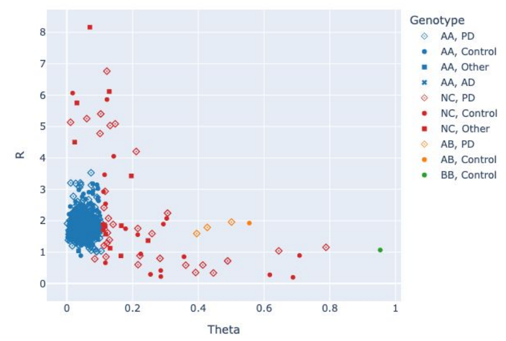

#### *GBA1* p.L424P

1:155235798:A:G GenTrain Score: 0.805194

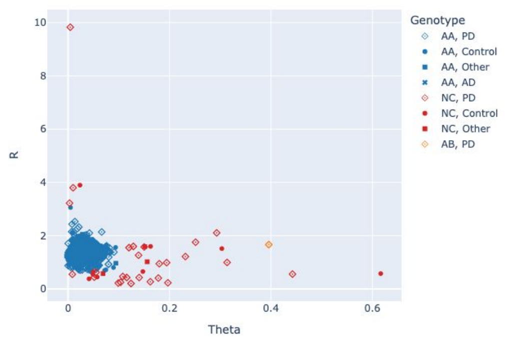

#### *GBA1* p.W420* (excluded)

1:155235810:C:T GenTrain Score: 0.851925

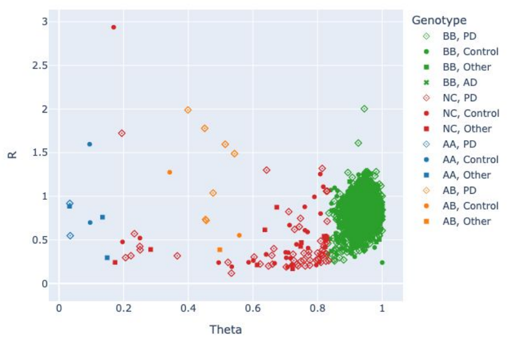

#### *GBA1* p.W417* (excluded)

1:155235819:C:T GenTrain Score: 0.866521

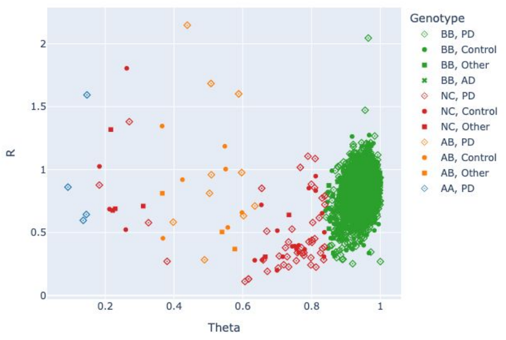

#### *GBA1* p.G416S

1:155235823:C:T GenTrain Score: 0.84641

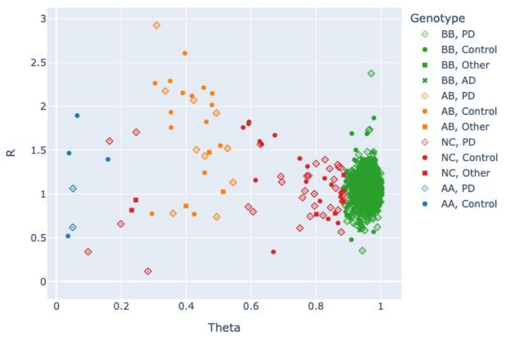

#### *GBA1* p.G416S

1:155235823:C:T GenTrain Score: 0.783854

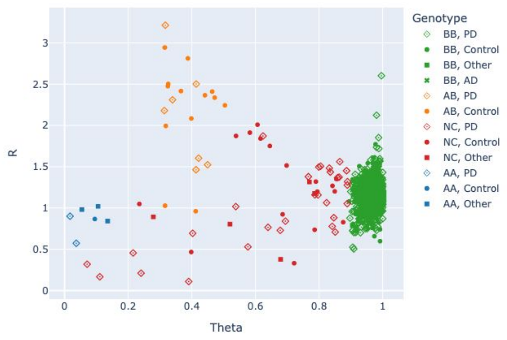

#### *GBA1* p.N409S

1:155235843:T:C GenTrain Score: 0.718304

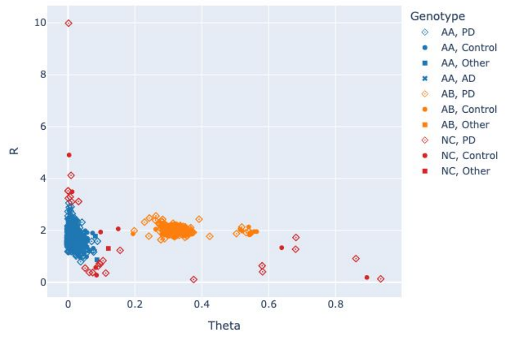

#### *GBA1* p.N409S

1:155235843:T:C GenTrain Score: 0.351873

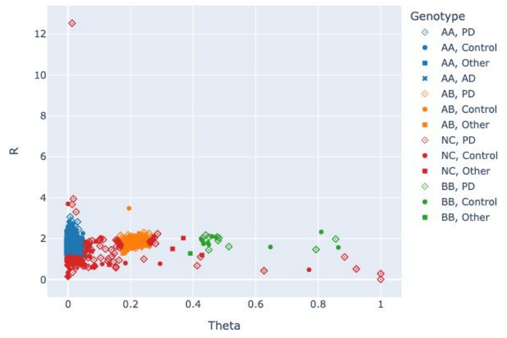

#### *GBA1* p.N409S

1:155235843:T:C GenTrain Score: 0.71119

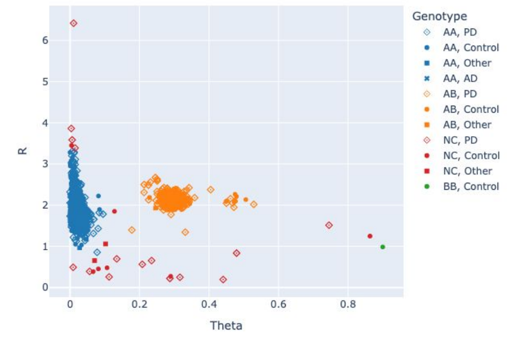

##### *GBA1* p.T408M

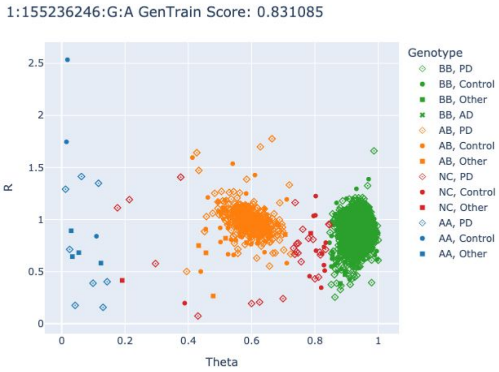

##### *GBA1* p.T408M

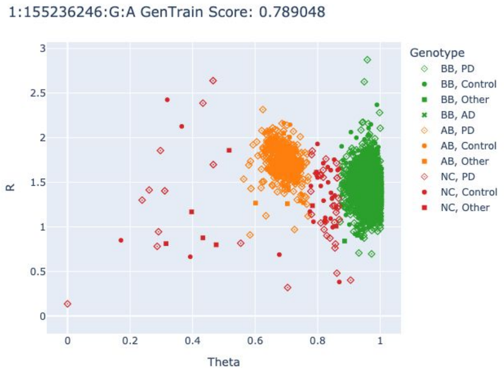

##### *GBA1* p.T408M

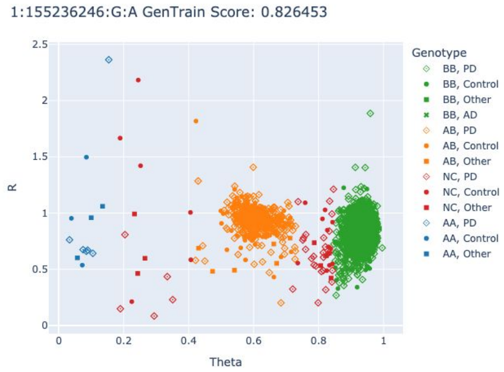

##### *GBA1* p.R368C (excluded)

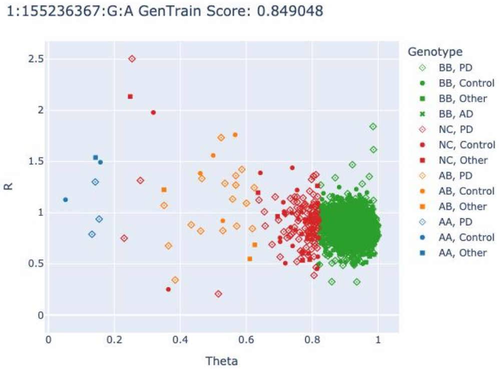

##### *GBA1* p.R368C (excluded)

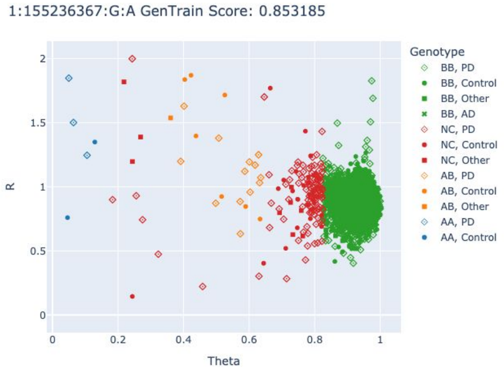

##### *GBA1* p.E365K

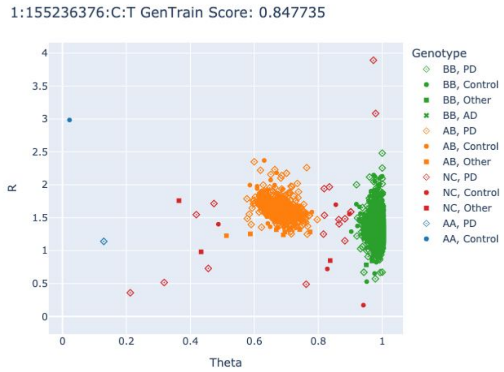

##### *GBA1* p.E365K

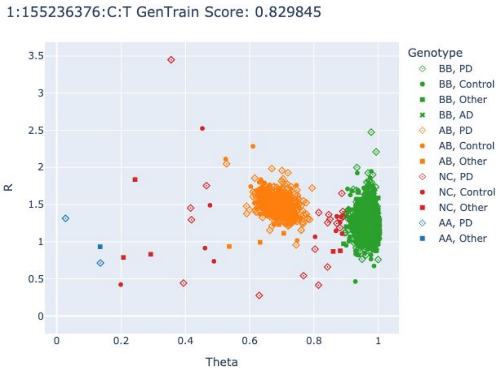

##### *GBA1* p.L363P

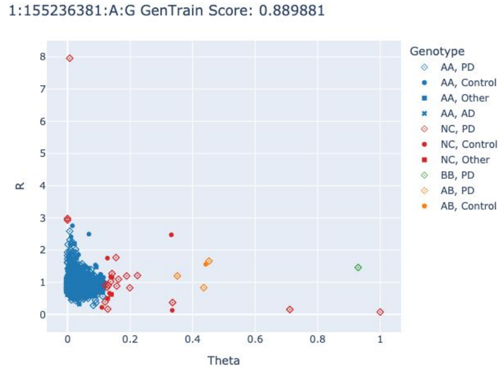

##### *GBA1* p.T362I

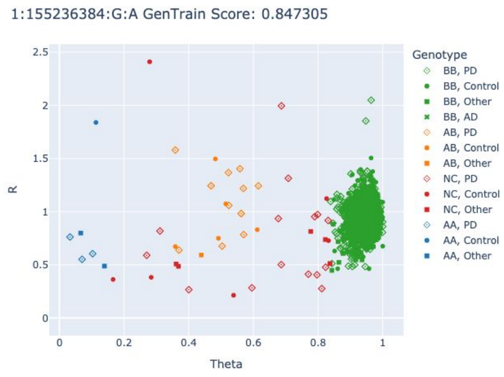

##### *GBA1* p.T362I

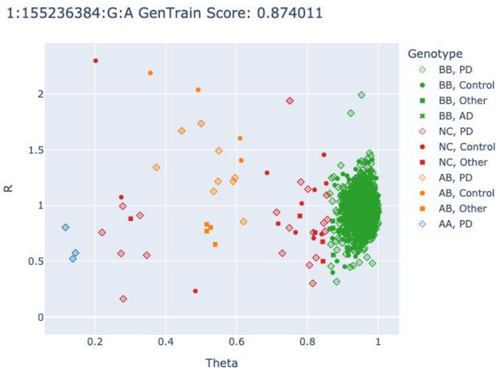

##### *GBA1* p.S310G

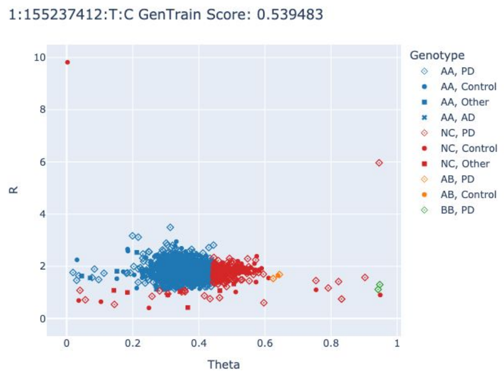

##### *GBA1* p.G241R

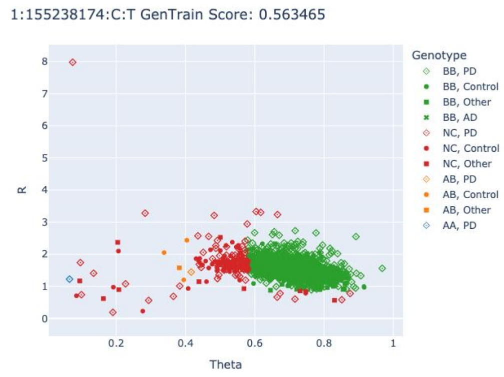

#### *GBA1* p.V230G

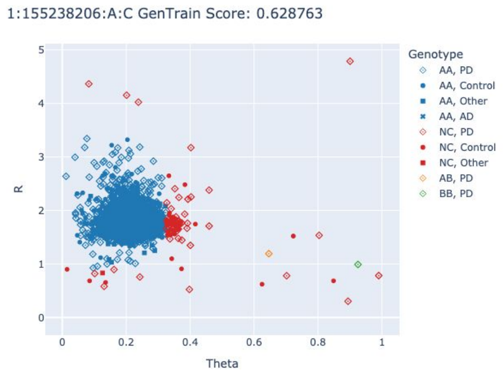

#### *GBA1* p.R296Q

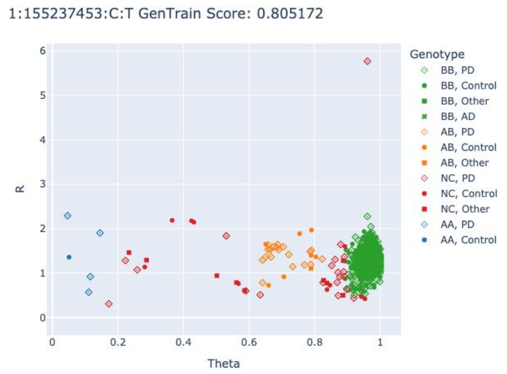

#### *GBA1* p.R296Q

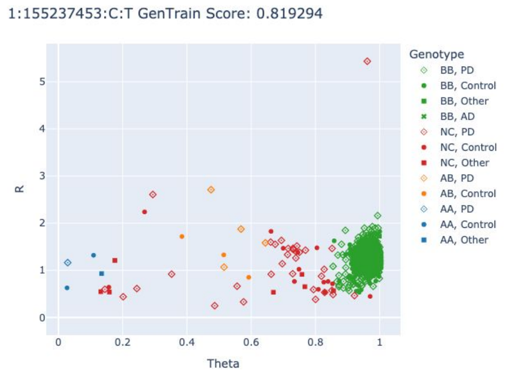

#### *GBA1* p.N227K

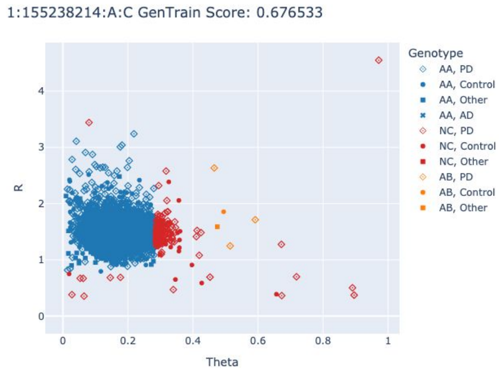

#### *GBA1* p.N227K

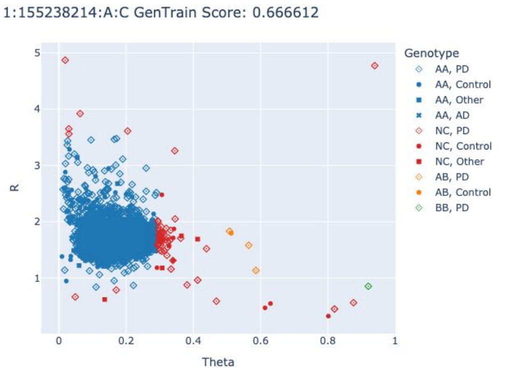

#### *GBA1* p.N227S

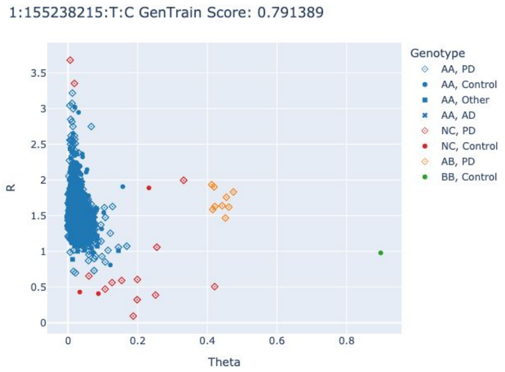

#### *GBA1* p.N227S

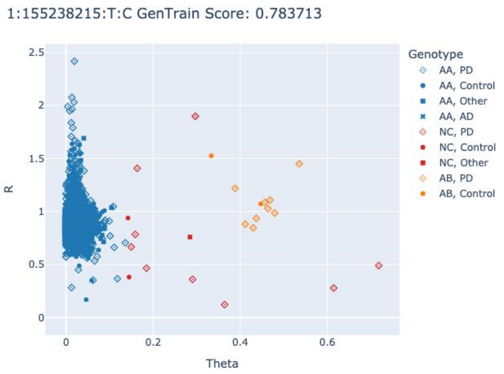

#### *GBA1* p.W223R (excluded)

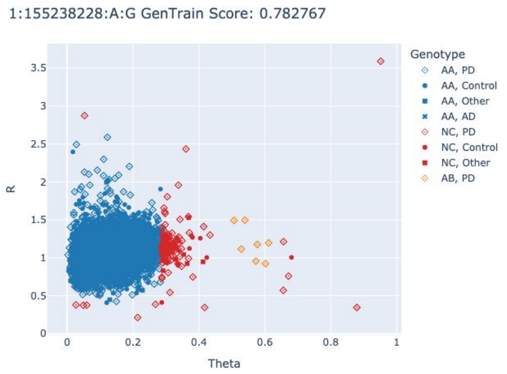

#### *GBA1* p.W218*

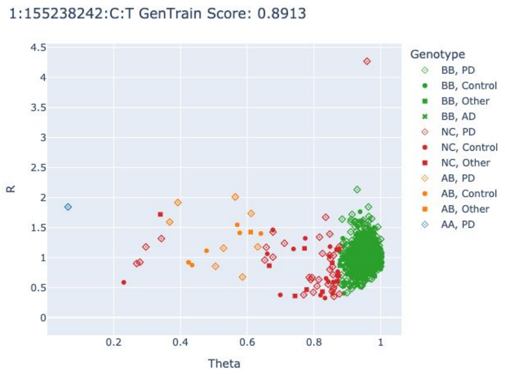

#### *GBA1* p.W218*

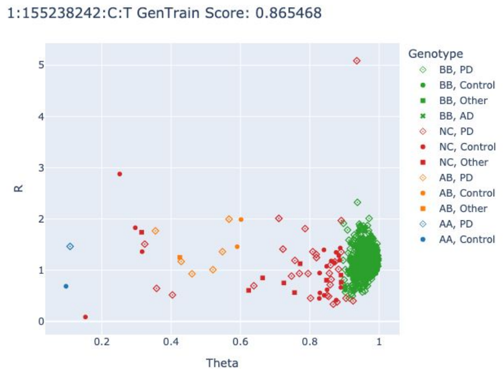

#### *GBA1* c.589-1G>A (excluded)

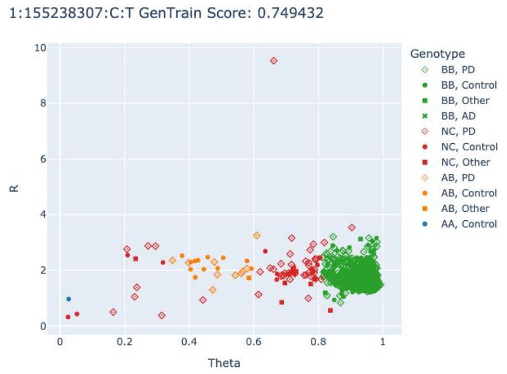

#### *GBA1* p.R159Q

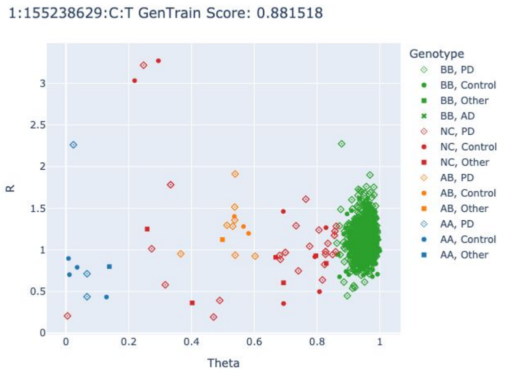

##### *GBA1* p.R159Q

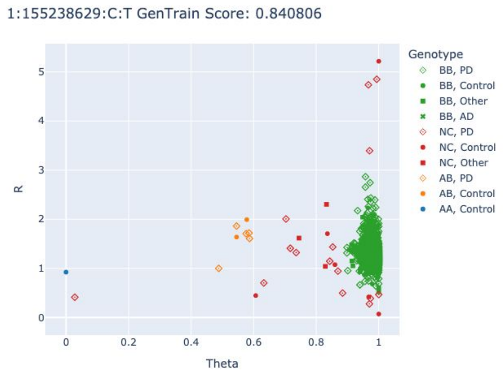

##### *GBA1* p.S146L

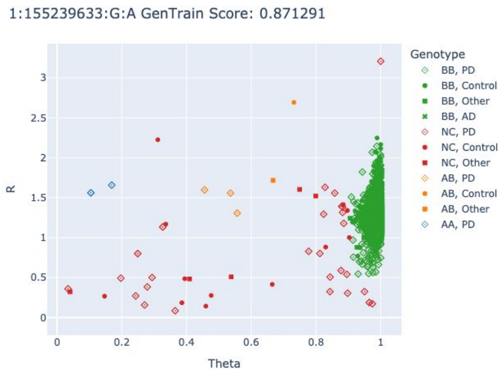

##### *GBA1* p.S146L

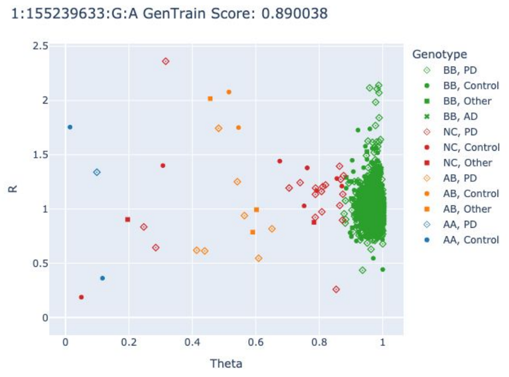

##### *GBA1* p.R87W

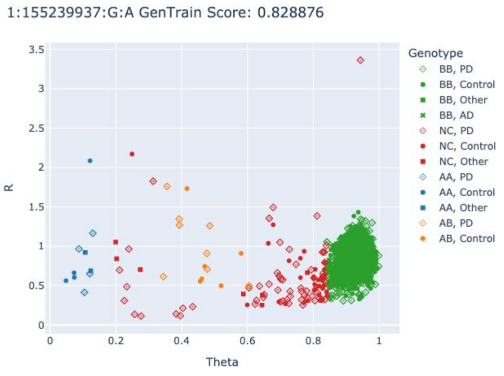

#### Cluster plots *SNCA* variants

##### *SNCA* p.A53T

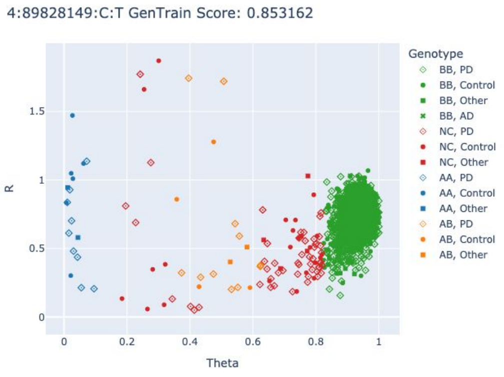

##### *SNCA* p.G51D

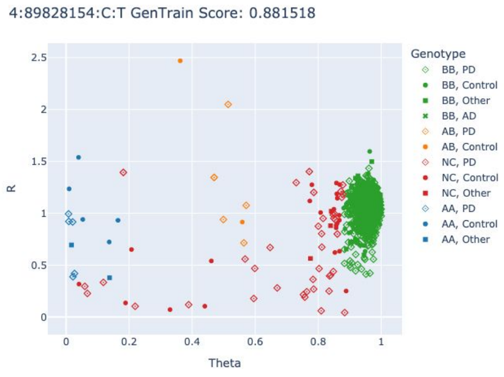

### Cluster plots *PRKN* variants

##### *PRKN* p.T415N

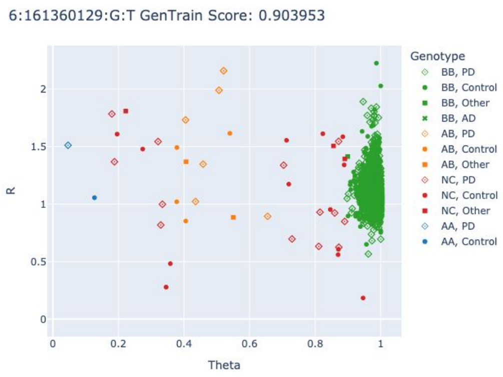

##### *PRKN* p.T415N

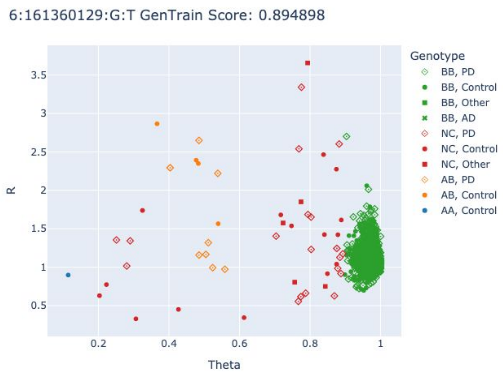

##### *PRKN* c.1083+1G>A

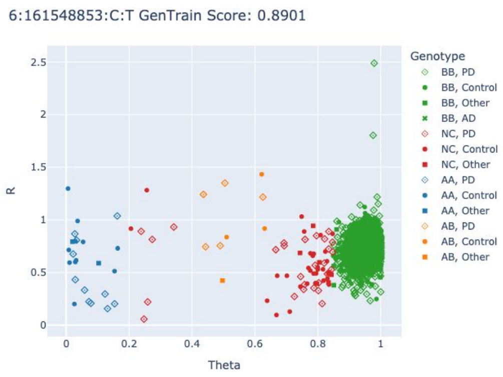

##### *PRKN* c.1083+1G>A

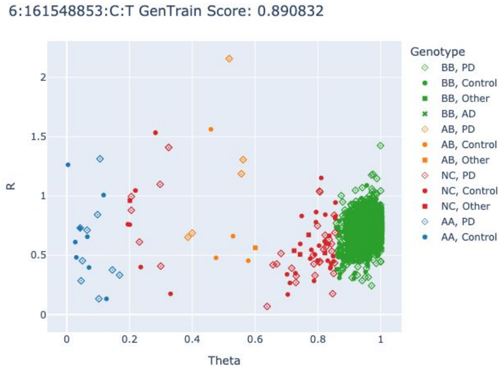

##### *PRKN* p.R275W

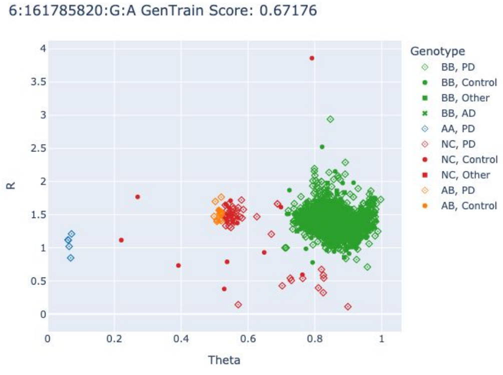

##### *PRKN* p.R275W

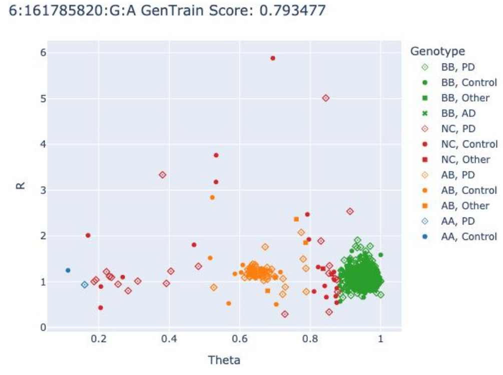

##### *PRKN* p.R275W

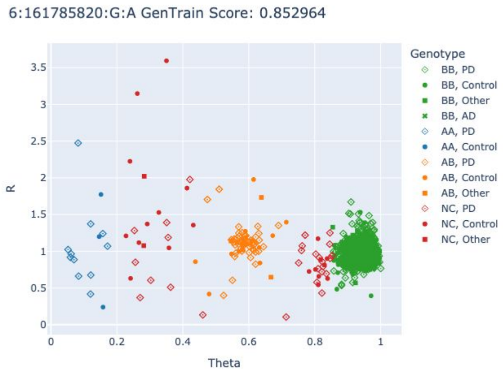

##### *PRKN* p.C253F

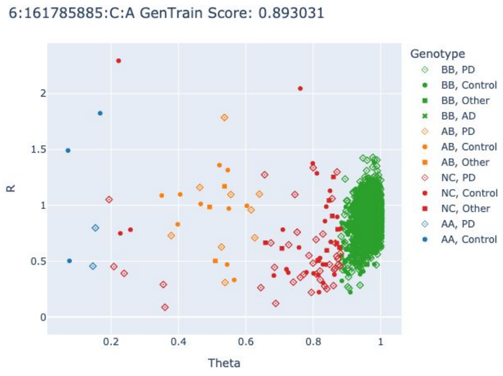

##### *PRKN* p.T240M

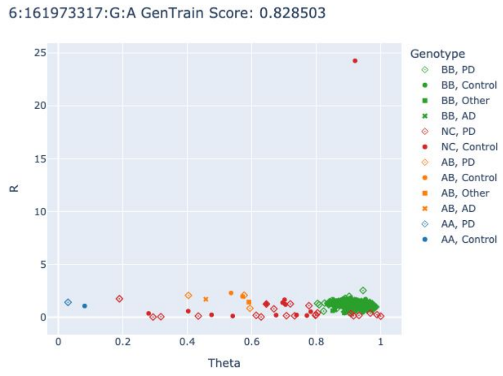

##### *PRKN* p.T240M

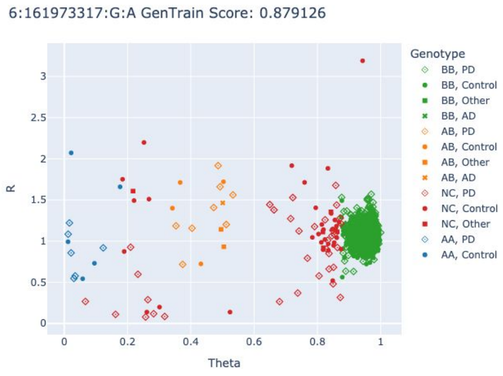

##### *PRKN* p.C238W

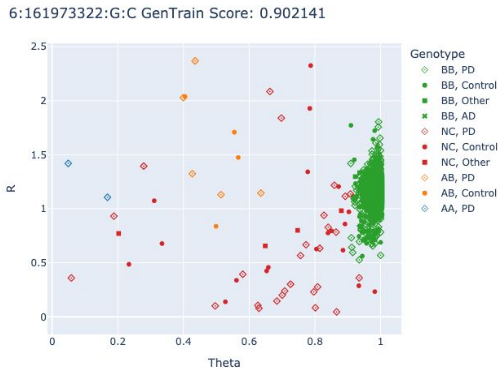

##### *PRKN* c.619-1G>A (excluded)

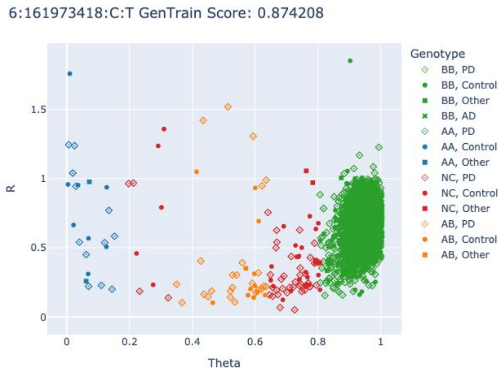

***PRKN* p.R33***

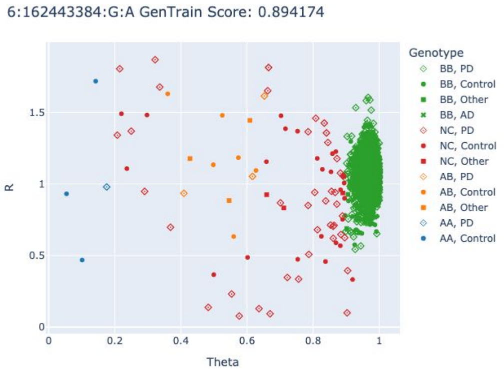

***PRKN* p.R33***

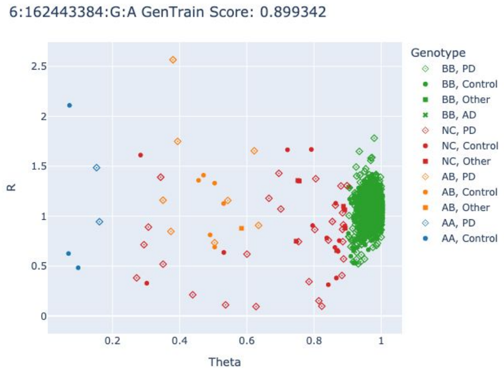

***PRKN* p.M1? (excluded)**

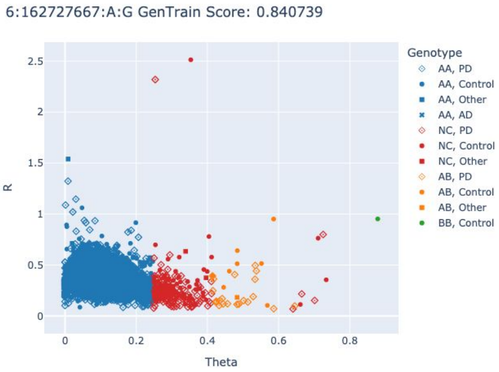

***PRKN* p.M1? (excluded)**

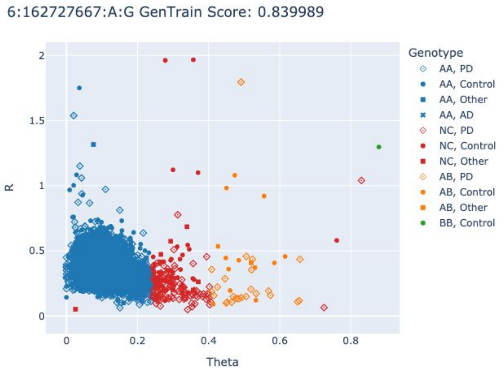

### Cluster plots *SLC20A2* variants

***SLC20A2* p.W551* (excluded)**

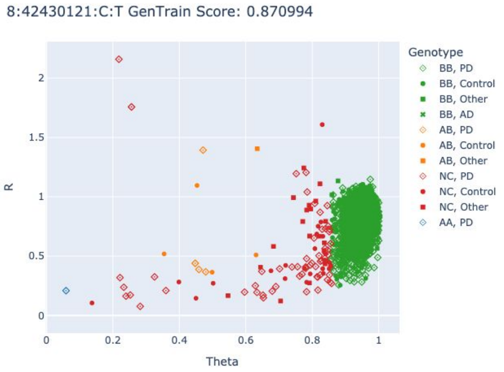

***SLC20A2* p.W551* (excluded)**

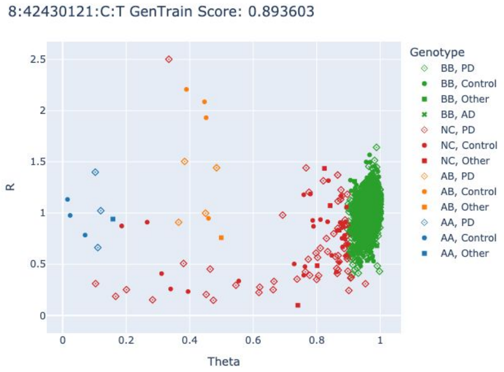

***SLC20A2* p.R254* (excluded)**

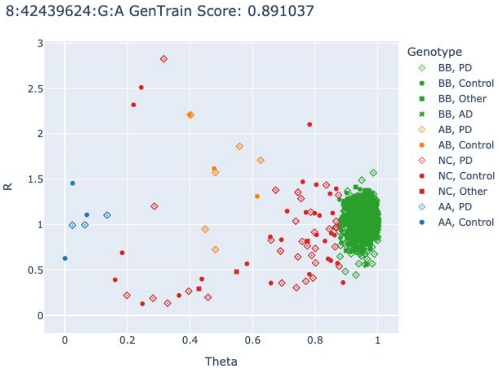

***SLC20A2* c.730+1G>A (excluded)**

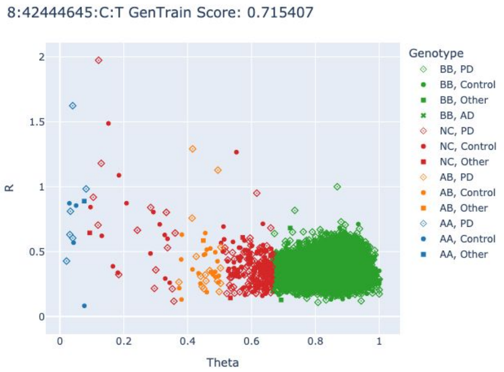

### Cluster plots *LRRK2* variants

#### *LRRK2* p.R1067Q

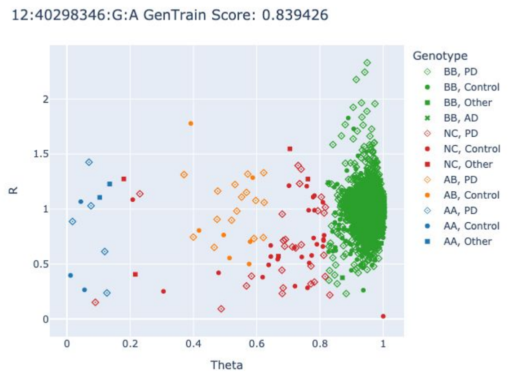

#### *LRRK2* p.R1067Q

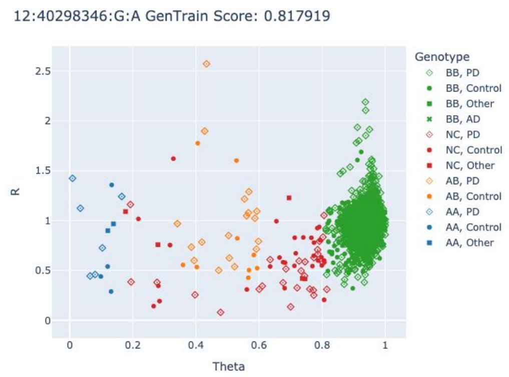

#### *LRRK2* p.R1325Q

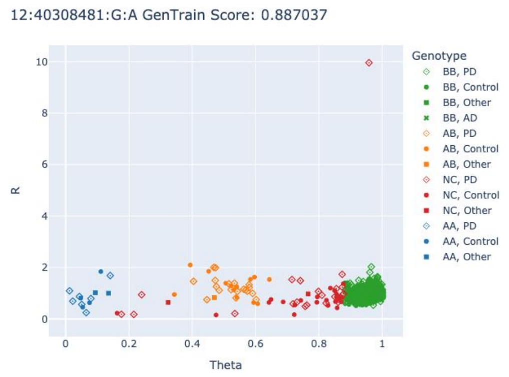

#### *LRRK2* p.R1325Q

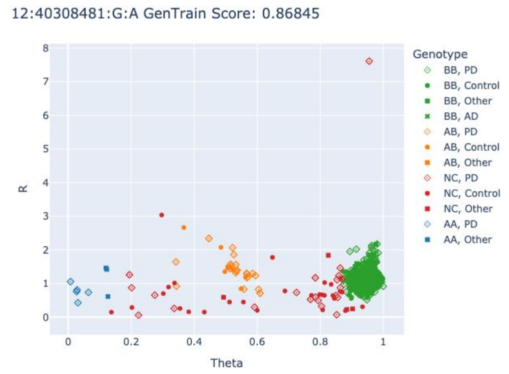

#### *LRRK2* p.R1325Q

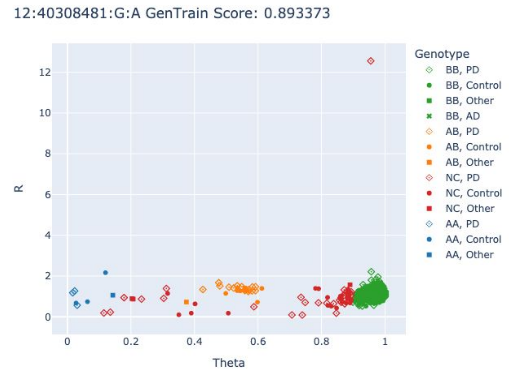

#### *LRRK2* p.R1441C

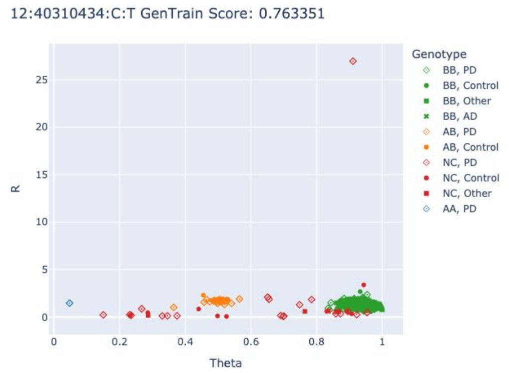

#### *LRRK2* p.R1441C

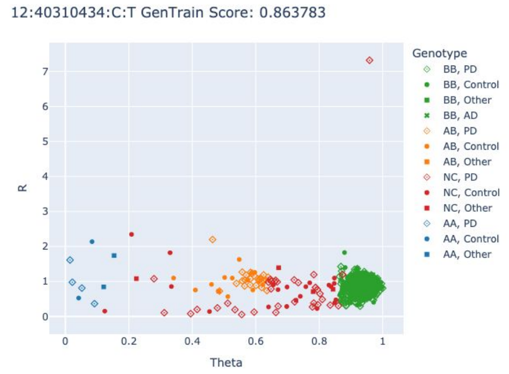

#### *LRRK2* p.R1441G

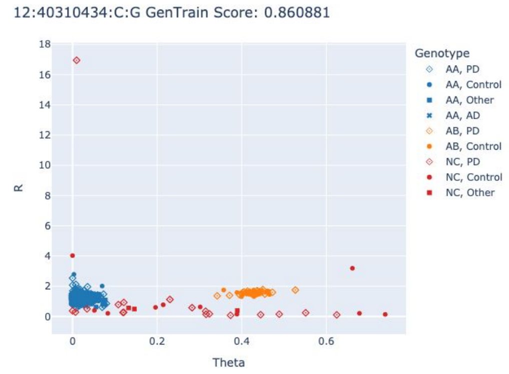

#### *LRRK2* p.R1441H

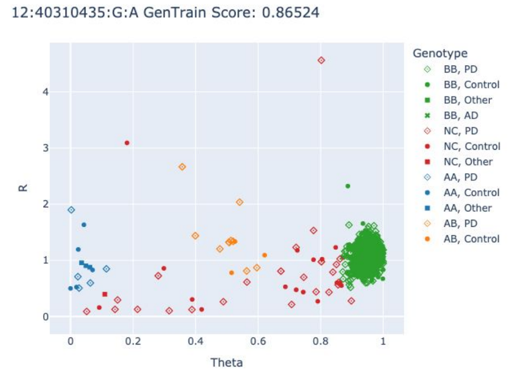

#### *LRRK2* p.R1441H

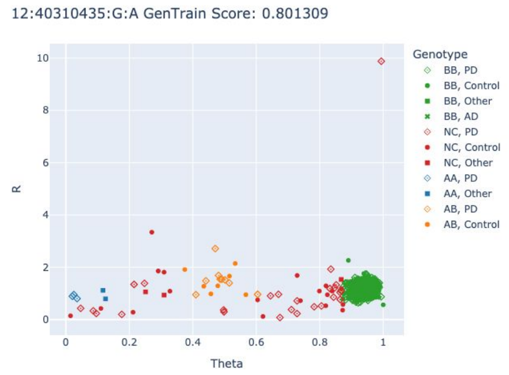

#### *LRRK2* p.R1628P

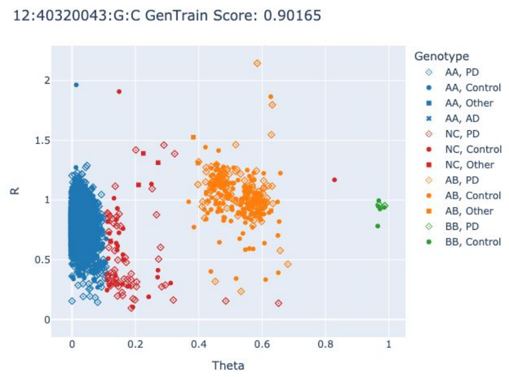

#### *LRRK2* p.Y1699C

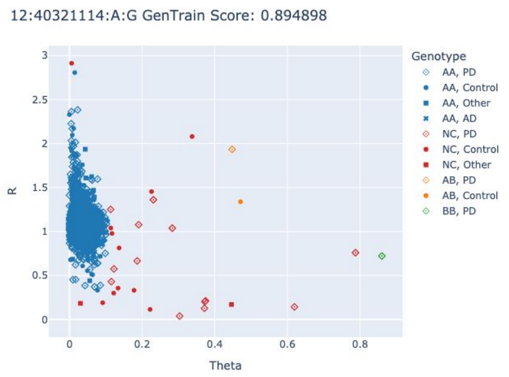

***LRRK2* p.Y1699C**

12:40321114:A:G GenTrain Score: 0.892127

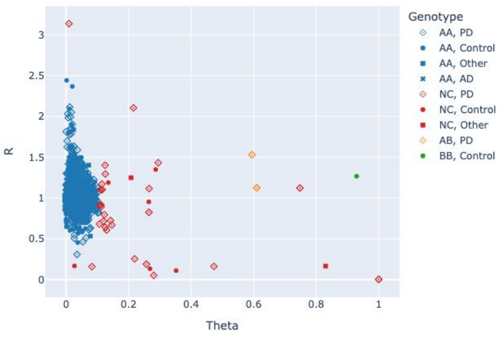

***LRRK2* p.L1795F**

12:40322386:G:T GenTrain Score: 0.838922

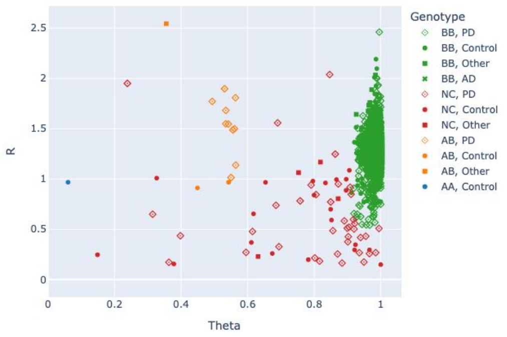

***LRRK2* p.L1795F**

12:40322386:G:T GenTrain Score: 0.880739

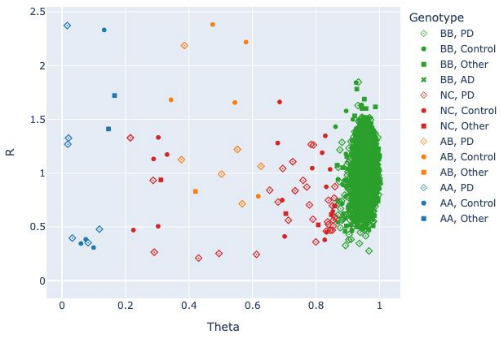

***LRRK2* p.G2019S**

12:40340400:G:A GenTrain Score: 0.842857

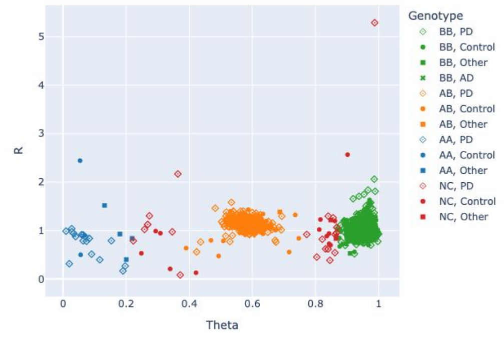

***LRRK2* p.G2019S**

12:40340400:G:A GenTrain Score: 0.814235

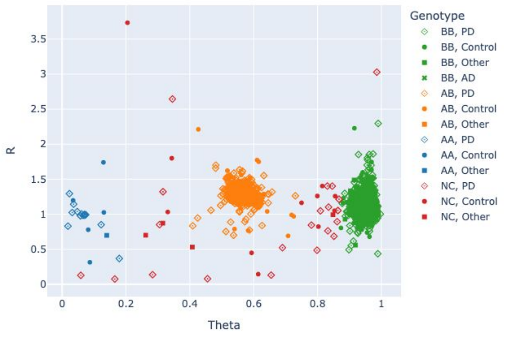

***LRRK2* p.G2019S**

12:40340400:G:A GenTrain Score: 0.833988

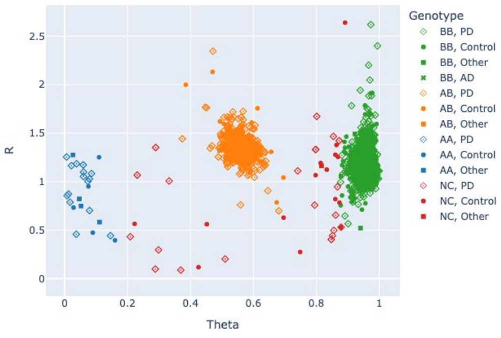

***LRRK2* p.I2020T**

12:40340404:T:C GenTrain Score: 0.892072

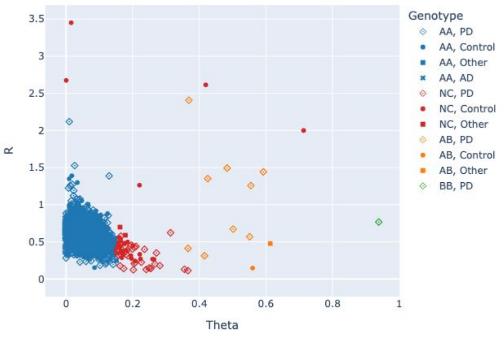

***LRRK2* p.I2020T**

12:40340404:T:C GenTrain Score: 0.8913

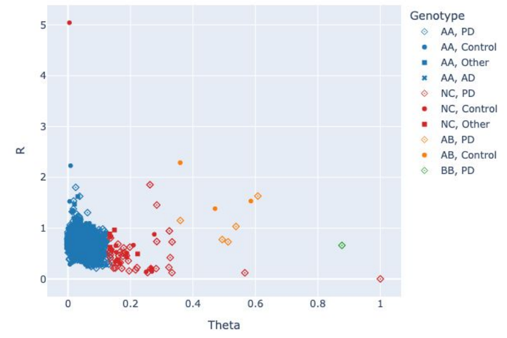

***LRRK2* p.G2385R**

12:40363526:G:A GenTrain Score: 0.8756

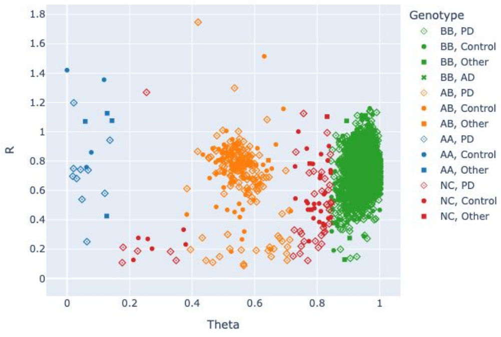

***LRRK2* p.G2385R**

12:40363526:G:A GenTrain Score: 0.889293

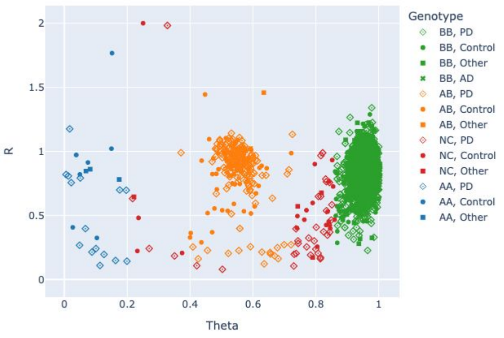

***LRRK2* p.G2385R**

12:40363526:G:A GenTrain Score: 0.87915

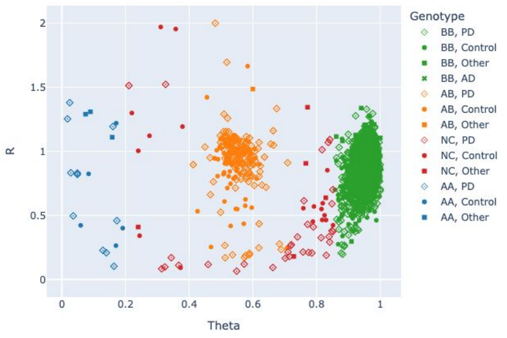

### Cluster plots *WDR45* variants

##### *WDR45* c.974-1C>T (excluded)

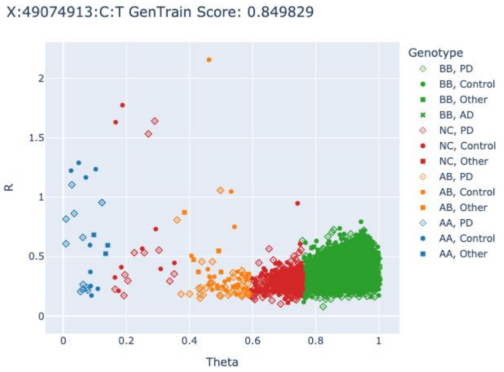

##### *WDR45* c.56-1G>A (excluded)

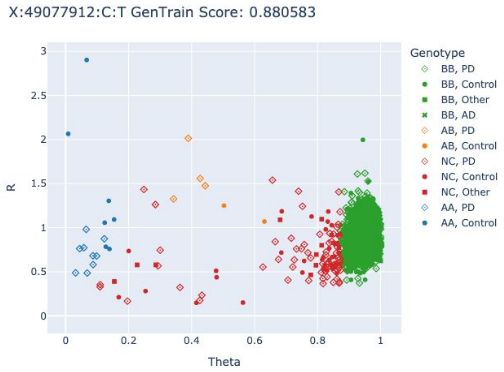

##### *WDR45* c.828-1G>C (excluded)

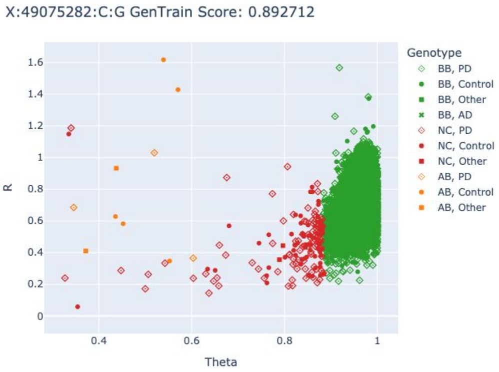

##### *WDR45* c.828-1G>C (excluded)

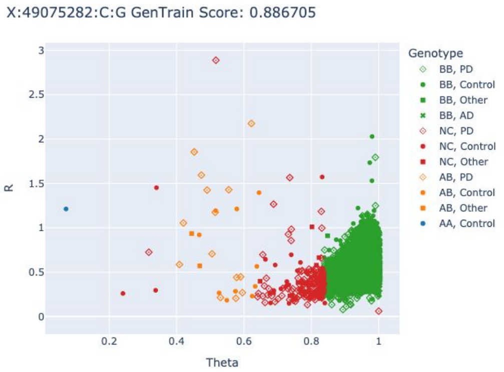

##### *WDR45* p.R233* (excluded)

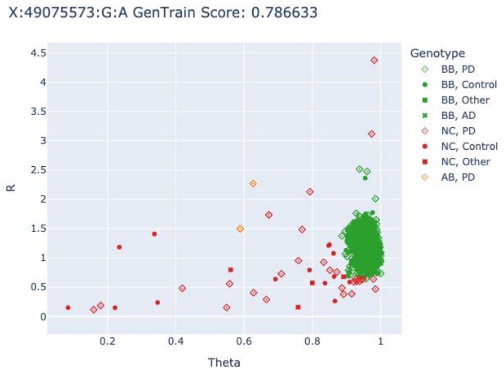

##### *WDR45* p.R233* (excluded)

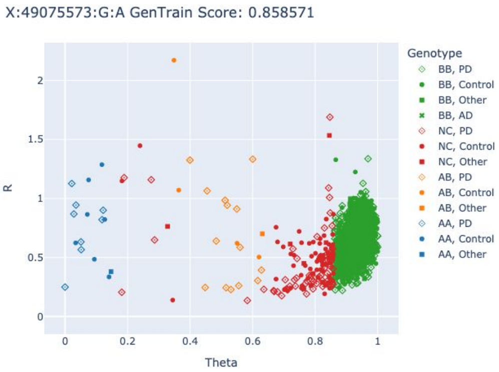

#### Supplementary Tables

**Supplementary Table 1. Overview of included samples investigated in this study.**

| | | AAC | AFR | AJ | AMR | CAH | CAS | EAS | EUR | FIN | MDE | SAS | **Total** |
|---|---|---|---|---|---|---|---|---|---|---|---|---|---|
| **NBA only** | **PD** | 183 | 1250 | 824 | 1627 | 412 | 535 | 1873 | 22562 | 78 | 363 | 234 | **29941** |
| | **Other phenotypes** | 7 | 11 | 50 | 18 | 8 | 19 | 187 | 1527 | 5 | 10 | 20 | **1862** |
| | **Controls** | 721 | 2352 | 645 | 1339 | 305 | 489 | 1751 | 24002 | 16 | 356 | 98 | **32074** |
| | **Healthy family members** | 1 | 0 | 0 | 0 | 0 | 12 | 4 | 37 | 0 | 1 | 1 | **56** |
| **WGS only** | **PD** | 61 | 74 | 473 | 574 | 102 | 95 | 187 | 5683 | 11 | 275 | 60 | **7595** |
| | **Other phenotypes** | 0 | 2 | 7 | 1 | 22 | 3 | 276 | 295 | 1 | 1 | 5 | **613** |
| | **Controls** | 35 | 82 | 109 | 146 | 35 | 107 | 18 | 1163 | 0 | 31 | 35 | **1761** |
| | **Healthy family members** | 0 | 1 | 0 | 0 | 1 | 13 | 12 | 68 | 0 | 1 | 0 | **96** |
| **NBA + CES** | **PD** | 121 | 74 | 641 | 263 | 235 | 28 | 86 | 4775 | 13 | 37 | 70 | **6343** |
| | **Other phenotypes** | 0 | 0 | 0 | 0 | 0 | 0 | 0 | 0 | 0 | 0 | 0 | **0** |
| | **Controls** | 0 | 0 | 0 | 0 | 0 | 0 | 0 | 0 | 0 | 0 | 0 | **0** |
| | **Healthy family members** | 0 | 0 | 0 | 0 | 0 | 0 | 0 | 0 | 0 | 0 | 0 | **0** |
| **NBA + WGS** | **PD** | 175 | 1444 | 367 | 321 | 138 | 624 | 2621 | 7458 | 22 | 671 | 431 | **14272** |
| | **Other phenotypes** | 4 | 30 | 41 | 7 | 13 | 109 | 199 | 2331 | 3 | 17 | 49 | **2803** |
| | **Controls** | 125 | 2061 | 159 | 169 | 104 | 884 | 1040 | 1596 | 6 | 953 | 307 | **7404** |
| | **Healthy family members** | 2 | 0 | 6 | 2 | 5 | 41 | 64 | 183 | 0 | 4 | 43 | **350** |
| **Other** | **PD** | 10 | 2 | 38 | 24 | 15 | 2 | 6 | 295 | 3 | 1 | 12 | **408** |
| | **Other phenotypes** | 0 | 0 | 0 | 0 | 0 | 0 | 0 | 0 | 0 | 0 | 0 | **0** |
| | **Controls** | 0 | 0 | 0 | 0 | 0 | 0 | 0 | 0 | 0 | 0 | 0 | **0** |
| | **Healthy family members** | 0 | 0 | 0 | 0 | 0 | 0 | 0 | 0 | 0 | 0 | 10 | **10** |
| **Total** | **PD** | 550 | 2844 | 2343 | 2809 | 902 | 1284 | 4773 | 40773 | 127 | 1347 | 807 | **58559** |
| | **Other phenotypes** | 11 | 43 | 98 | 26 | 43 | 131 | 662 | 4153 | 9 | 28 | 74 | **5278** |
| | **Controls** | 881 | 4495 | 913 | 1654 | 444 | 1480 | 2809 | 26761 | 22 | 1340 | 440 | **41239** |
| | **Healthy family members** | 3 | 1 | 6 | 2 | 6 | 66 | 80 | 288 | 0 | 6 | 54 | **512** |
| **TOTAL** | **Total unique samples** | **1445** | **7383** | **3360** | **4491** | **1395** | **2961** | **8324** | **71975** | **158** | **2721** | **1375** | **105588** |

The "Affected" group summarizes the number of samples of all affected individuals investigated in this study, including individuals from unselected PD cohorts, individuals submitted to the monogenic GP2 study arm (e.g., samples with an AAO ≤50 years and/or a positive family history of PD), individuals with other neurological and neurodegenerative phenotypes (e.g., atypical parkinsonism, different types of dementia, SWEDD, tremor, etc.) and affected individuals submitted as part of genetically enriched cohorts (e.g., known *GBA1* or *LRRK2* variant carriers). The "Unaffected" group summarizes the number of samples of all unaffected individuals investigated in this study, including healthy controls, unaffected family members of individuals with PD, individuals from population cohorts, and unaffected individuals submitted as part of genetically enriched cohorts (e.g., asymptomatic *GBA1* and *LRRK2* carrier).

AAC = African Admixed, AFR = African, AJ = Ashkenazi Jewish, AMR = Latino and Indigenous people of the Americas, CAH = Complex Admixture, CAS = Central Asian, CES = Clinical exome sequencing, EAS = East Asian, EUR = European, FIN = Finnish, MDE = Middle Eastern, NBA = NeuroBooster Array (genotyping), SAS = South Asian, WGS = short-read whole genome sequencing.

**Supplementary Table 2. Summary of genetic findings across all ancestries.**

| Genetic ancestry | Group | Investigated samples (n) | PD risk* | | Typical autosomal-dominant PD | | | | Early-onset recessive PD | | | Atypical parkinsonism | *Dual carriers*** | Total (%) |
|---|---|---|---|---|---|---|---|---|---|---|---|---|---|---|
| | | | *GBA1* | *LRRK2* | *LRRK2* | *SNCA* | *VPS35* | *RAB32* | *PINK1* | *PRKN* | *PARK7* | | | |
| **AAC** | PD | 550 | 171 / 3 / 24 | 0 | 1 | 0 | 0 | 0 | 0 | 1 | 0 | 1 | 2 | 199/550 (36.2) |
| | Other phenotypes | 11 | 2 / 0 / 0 | 0 | 0 | 0 | 0 | 0 | 0 | 0 | 0 | 0 | 0 | 2/11 (18.2) |
| | Controls | 881 | 201 / 3 / 6 | 1 | 3 | 0 | 0 | 0 | 0 | 0 | 0 | 0 | 1 | 213/881 (24.2) |
| | Healthy family members | 3 | 0 | 0 | 1 | 0 | 0 | 0 | 0 | 0 | 0 | 0 | 0 | 1/3 (33.3) |
| **AFR** | PD | 2844 | 1133 / 33 / 339 | 2 | 3 | 2 | 0 | 0 | 1 | 4 | 0 | 0 | 7 | 1510/2844 (53.1) |
| | Other phenotypes | 43 | 11 / 1 / 2 | 0 | 0 | 0 | 0 | 0 | 0 | 0 | 0 | 0 | 0 | 14/43 (32.6) |
| | Controls | 4495 | 1504 / 3 / 184 | 1 | 3 | 0 | 0 | 0 | 0 | 0 | 0 | 1 | 3 | 1693/4495 (37.7) |
| | Healthy family members | 1 | 0 | 0 | 0 | 0 | 0 | 0 | 0 | 0 | 0 | 0 | 0 | 0/1 (0) |
| **AJ** | PD | 2343 | 383 / 6 / 5 | 0 | 245 / 5 | 1 | 0 | 0 | 0 | 0 | 0 | 0 | 25 | 620/2343 (26.5) |
| | Other phenotypes | 98 | 15 / 0 / 0 | 0 | 2 | 0 | 0 | 0 | 0 | 0 | 0 | 0 | 1 | 16/98 (16.3) |
| | Controls | 913 | 73 / 1 / 2 | 0 | 20 | 0 | 0 | 0 | 0 | 0 | 0 | 0 | 1 | 95/913 (10.4) |
| | Healthy family members | 6 | 0 | 0 | 2 | 0 | 0 | 0 | 0 | 0 | 0 | 0 | 0 | 2/6 (33.3) |
| **AMR** | PD | 2809 | 151 / 8 / 3 | 0 | 59 | 2 | 0 | 0 | 0 | 24 | 0 | 0 | 2 | 245/2809 (8.7) |
| | Other phenotypes | 26 | 0 / 0 / 0 | 0 | 0 | 0 | 0 | 0 | 0 | 0 | 0 | 2 | 0 | 2/26 (7.7) |
| | Controls | 1654 | 30 / 0 / 0 | 0 | 0 | 0 | 0 | 0 | 0 | 0 | 0 | 0 | 0 | 30/1654 (1.8) |
| | Healthy family members | 2 | 0 / 0 / 1 | 0 | 0 | 0 | 0 | 0 | 0 | 0 | 0 | 0 | 0 | 1/2 (50.0) |
| **CAH** | PD | 902 | 130 / 2 / 7 | 9 | 21 | 1 | 1 | 0 | 0 | 12 | 0 | 0 | 4 | 179/902 (19.8) |
| | Other phenotypes | 43 | 2 / 0 / 0 | 1 | 0 | 0 | 0 | 0 | 0 | 0 | 0 | 0 | 0 | 3/43 (7.0) |
| | Controls | 444 | 40 / 1 / 2 | 3 | 2 | 0 | 0 | 0 | 0 | 0 | 0 | 0 | 0 | 48/444 (10.8) |
| | Healthy family members | 6 | 0 | 0 | 3 | 0 | 0 | 0 | 0 | 0 | 0 | 0 | 0 | 3/6 (50) |
| **CAS** | PD | 1284 | 73 / 1 / 1 | 23 | 1 | 0 | 0 | 1 | 0 | 7 | 0 | 0 | 2 | 105/1284 (8.2) |
| | Other phenotypes | 131 | 1 / 0 / 0 | 1 | 0 | 0 | 0 | 0 | 0 | 0 | 0 | 0 | 0 | 2/131 (1.5) |
| | Controls | 1480 | 29 / 0 / 0 | 14 | 0 | 0 | 0 | 0 | 0 | 1 | 0 | 0 | 0 | 44/1480 (3.0) |
| | Healthy family members | 66 | 1 / 0 / 0 | 0 | 0 | 0 | 0 | 0 | 0 | 0 | 0 | 0 | 0 | 1/66 (1.5) |
| **EAS** | PD | 4773 | 190 / 5 / 0 | 574 / 16 / 11 | 15 | 4 | 5 | 0 | 17 | 17 | 0 | 1 | 25 | 840/4773 (17.6) |
| | Other phenotypes | 662 | 9 / 0 / 0 | 49 / 1 / 1 | 1 | 0 | 0 | 0 | 0 | 0 | 0 | 0 | 1 | 60/662 (9.1) |
| | Controls | 2809 | 14 / 1 / 0 | 165 / 4 / 1 | 2 | 0 | 0 | 0 | 0 | 0 | 0 | 0 | 0 | 187/2809 (6.7) |
| | Healthy family members | 80 | 2 / 0 / 0 | 7 / 0 / 3 | 0 | 0 | 0 | 0 | 0 | 0 | 0 | 0 | 0 | 12/80 (15.0) |
| **EUR** | PD | 40773 | 3312 / 99 / 20 | 21 | 451 / 2 | 31 | 8 | 17 | 5 | 126 | 3 | 8 | 49 | 4054/40773 (9.9) |
| | Other phenotypes | 4153 | 241 / 7 / 1 | 4 | 8 | 0 | 0 | 0 | 0 | 0 | 0 | 1 | 0 | 262/4153 (6.3) |

| | | | | | | | | | | | | | | |
|---|---|---|---|---|---|---|---|---|---|---|---|---|---|---|
| | Controls | 26761 | 1255 / 11 / 2 | 6 | 65 | 0 | 0 | 0 | 0 | 2 | 0 | 2 | 6 | 1337/26761 (6.25) |
| | Healthy family members | 288 | 23 / 2 / 0 | 0 | 4 | 0 | 0 | 0 | 0 | 0 | 0 | 0 | 0 | 29/288 (10.1) |
| **FIN** | PD | 127 | 25 / 1 / 0 | 0 | 0 | 0 | 0 | 0 | 0 | 1 | 0 | 0 | 1 | 26/127 (20.5) |
| | Other phenotypes | 9 | 3 / 0 / 0 | 0 | 0 | 0 | 0 | 0 | 0 | 0 | 0 | 0 | 0 | 3/9 (33.3) |
| | Controls | 22 | 4 / 0 / 0 | 0 | 0 | 0 | 0 | 0 | 0 | 0 | 0 | 0 | 0 | 4/22 (18.2) |
| | Healthy family members | 0 | 0 | 0 | 0 | 0 | 0 | 0 | 0 | 0 | 0 | 0 | 0 | 0/0 (0) |
| **MDE** | PD | 1347 | 70 / 1 / 2 | 0 | 56 / 3 | 1 | 0 | 4 | 11 | 17 | 0 | 0 | 7 | 158/1347 (11.7) |
| | Other phenotypes | 28 | 2 / 0 / 0 | 0 | 1 | 0 | 0 | 0 | 0 | 0 | 0 | 0 | 0 | 3/28 (10.7) |
| | Controls | 1340 | 20 / 0 / 0 | 0 | 4 | 0 | 0 | 0 | 0 | 1 | 0 | 0 | 0 | 25/1340 (1.9) |
| | Healthy family members | 6 | 0 | 0 | 0 | 0 | 0 | 0 | 0 | 0 | 0 | 0 | 0 | 0/6 (0) |
| **SAS** | PD | 807 | 38 / 0 / 1 | 0 | 1 | 0 | 1 | 0 | 4 | 5 | 0 | 0 | 1 | 49/807 (6.1) |
| | Other phenotypes | 74 | 1 / 0 / 0 | 0 | 0 | 0 | 0 | 0 | 0 | 0 | 0 | 0 | 0 | 1/74 (1.4) |
| | Controls | 440 | 4 / 0 / 0 | 0 | 0 | 0 | 0 | 0 | 0 | 0 | 0 | 0 | 0 | 4/440 (0.9) |
| | Healthy family members | 54 | 5 / 0 / 0 | 0 | 0 | 0 | 0 | 0 | 0 | 0 | 0 | 0 | 0 | 5/54 (9.3) |

This table reports gene-specific numbers of variant carriers across ancestries, with the total reflecting the number of unique individuals. For autosomal dominant genes, counts reflect heterozygous carriers, whereas for autosomal recessive genes, counts reflect biallelic carriers. In columns with three different values, the first denotes heterozygous carriers, the second individuals carrying two different heterozygous variants, and the third homozygous variant carriers. For columns with two different values, the first denotes heterozygous and the second homozygous carriers.

* All pathogenic/likely pathogenic GBA1 variants are considered risk variants in the context of PD, including Gaucher disease-causing variants of all severities (e.g., severe and mild) as well as variants only associated with an increased risk of PD
(rs2230288, p.E365K [1:155236376:C:T]; rs75548401, p.T408M [1:155236246:G:A], and rs3115534-G [1:155235878:G:T]). LRRK2 PD risk variants include rs33949390 (chr12:40320043:G:C, p.R1628P) and rs34778348 (chr12:40363526:G:A, p.G2385R).

** Dual carriers refer to individuals harboring pathogenic/likely pathogenic or Parkinson’s disease risk variants in two different genes. The total number reflects the number of individuals rather than variants; therefore, dual carriers are counted only once in the total, and column-wise sums may exceed the total.

AAC = African admixed, AFR = African, AJ = Ashkenazi Jewish, AMR = Latinos and Indigenous people of the Americas, CAH = Complex admixture, CAS = Central Asian, EAS = East Asian, EUR = European, FIN = Finnish, MDE = Middle East, SAS = South Asian

**Supplementary Table 3. Summary of genetic findings across all ancestries in atypical parkinsonism genes.**

| Genetic ancestry | Group | Investigated samples (n) | Atypical parkinsonism | | | | Total (%) |
|---|---|---|---|---|---|---|---|
| | | | *ATP13A2* | *SLC20A2* | *RAB39B* | *WDR45* | |
| **AAC** | PD | 550 | 0 | 0 | 0 | 1 | 1/550 (0.2) |
| **AFR** | Controls | 4495 | 0 | 0 | 0 | 1 | 1/4495 (<0.1) |
| **AMR** | Other phenotypes | 26 | 2 | 0 | 0 | 0 | 2/26 (7.7) |
| **EAS** | PD | 4773 | 0 | 1 | 0 | 0 | 1/4773 (<0.1) |
| **EUR** | PD | 40773 | 0 | 1 | 2 | 5 | 8/40773 (<0.1) |
| | Other phenotypes | 4153 | 0 | 1 | 0 | 0 | 1/4153 (<0.1) |
| | Controls | 26761 | 0 | 0 | 0 | 2 | 2/26761 (<0.1) |

AAC = African admixed, AFR = African, AMR = Latinos and Indigenous people of the Americas, EAS = East Asian, EUR = European

**Supplementary Table 4. Results of linear regression analysis comparing age at onset (AAO) between idiopathic PD (IPD, reference group) and *LRRK2*- and *GBA1*-associated PD across ancestries.**

| Ancestry | Group | N | Coef. | Std.Err. | t | P | [0.025 | 0.975] |
|---|---|---|---|---|---|---|---|---|
| AAC | *GBA1*-PD compared to IPD | 541 | -3.442097412 | 1.309651559 | -2.628254354 | **0.008919635** | -6.016917047 | -0.867277778 |
| AFR | *GBA1*-PD compared to IPD | 2826 | -1.377408093 | 0.404961994 | -3.401326821 | **0.000680559** | -2.171482523 | -0.583333662 |
| AJ | *GBA1*-PD compared to IPD | 2302 | -3.657626015 | 0.739083914 | -4.948864324 | **8.14E-07** | -5.107156333 | -2.208095697 |
| | *LRRK2*-PD compared to IPD | 2302 | -2.974299693 | 0.916860394 | -3.244004989 | **0.001199721** | -4.772494611 | -1.176104774 |
| AMR | *GBA1*-PD compared to IPD | 2723 | -2.172395567 | 1.513659233 | -1.435194606 | 0.151521984 | -5.142450921 | 0.797659787 |
| | *LRRK2*-PD compared to IPD | 2723 | -1.121344156 | 2.867367466 | -0.391070963 | 0.695822133 | -6.747603956 | 4.504915645 |
| CAS | *GBA1*-PD compared to IPD | 1255 | -5.399863354 | 2.245605672 | -2.404635605 | **0.016493096** | -9.810140778 | -0.98958593 |
| | *LRRK2*-PD compared to IPD | 1255 | 2.188631945 | 4.408176402 | 0.49649373 | 0.619729481 | -6.468845683 | 10.84610957 |
| EAS | *GBA1*-PD compared to IPD | 4623 | -6.44542234 | 1.046418585 | -6.159506752 | **8.11E-10** | -8.497062221 | -4.393782459 |
| | *LRRK2*-PD compared to IPD | 4623 | -1.108071119 | 0.619113152 | -1.789771572 | 0.073575559 | -2.321923063 | 0.105780825 |
| EUR | *GBA1*-PD compared to IPD | 39727 | -1.670782364 | 0.246063789 | -6.790037524 | **1.14E-11** | -2.153077179 | -1.188487549 |
| | *LRRK2*-PD compared to IPD | 39727 | -1.208832136 | 0.64885302 | -1.863029222 | 0.062467514 | -2.480609864 | 0.062945591 |
| FIN | *GBA1*-PD compared to IPD | 126 | -5.453731793 | 3.099820108 | -1.759370416 | 0.083369764 | -11.64822799 | 0.740764401 |
| MDE | *GBA1*-PD compared to IPD | 1275 | -2.500207604 | 1.842298514 | -1.357113185 | 0.175124688 | -6.116469985 | 1.116054778 |
| | *LRRK2*-PD compared to IPD | 1275 | -2.699389161 | 1.841971719 | -1.465488928 | 0.143177066 | -6.315010076 | 0.916231754 |
| SAS | *GBA1*-PD compared to IPD | 784 | -8.021536914 | 2.127215908 | -3.770908671 | **0.000176617** | -12.19814029 | -3.844933541 |

Columns indicate the number of individuals included (N), regression coefficient (Coef.) representing the estimated AAO difference compared to IPD, standard error (Std.Err.), t-statistic (t), p-value (P), and 95% confidence interval ([0.025, 0.975]).
